# Acute-Phase Machine Learning Prediction of 12-Month Aphasia and Discourse Recovery

**DOI:** 10.64898/2026.05.13.26353123

**Authors:** Manuel Jose Marte, Mathew Chaves, Lindsey Kelly, Isidora Diaz-Carr, Voss Neal, Andreia V. Faria, Melissa D. Stockbridge, Argye E. Hillis

**Author notes:** **Corresponding author**: Argye E. Hillis, 600 N. Wolfe Street / Phipps 4-448 Baltimore, MD 21287-7613, T, F.

## Abstract

Approximately 30-40% of stroke patients retain aphasia at 12 months. Early forecasting may guide rehabilitation and prognostic enrichment of clinical trials, yet machine learning (ML) prediction of language recovery has typically relied on chronic-phase data unavailable at the acute decision point. Whether acute features predict 12-month outcomes, and whether global severity and connected-speech recovery share substrates in an ML framework, is untested. We studied 73 patients with acute left-hemisphere ischemic stroke and aphasia (mean 2.8 days post-onset). Two 12-month outcomes were defined: aphasia resolution (Western Aphasia Battery–Revised Aphasia Quotient [WAB-AQ] ≥93.8) and discourse normalization (Modern Cookie Theft content units ≥22.1; N=61). Four ML algorithms were trained on four hierarchical feature sets (clinical, volumetric, anatomical, network-disconnection) using nested cross-validation and SHapley Additive exPlanations (SHAP) stability analysis. Acute WAB-AQ dominated (mean |SHAP| = 13.60, ∼20× the next feature). For aphasia resolution, random forest achieved F1 = 0.874 (95% CI, 0.800–0.941), Pearson r = 0.827, mean absolute error (MAE) = 7.26 WAB-AQ points; clinical features alone achieved F1 = 0.851. For discourse, support vector regression achieved F1 = 0.725 (95% CI 0.593–0.831), r = 0.617, MAE = 8.96 content units. Three predictors were shared (acute WAB-AQ, lesion volume, left pars triangularis); ventral-stream tracts were linked to aphasia resolution, whereas interhemispheric and prefrontal connectivity were linked to discourse. Both models overpredicted severe chronic outcomes. Acute-phase ML forecasts 12-month aphasia resolution accurately and discourse more modestly. Clinical features carry most predictive variance; acute imaging reveals shared and outcome-specific substrates mapping onto dual-stream architecture, supporting early stratification for rehabilitation and prognostic trial enrichment.

**Abbreviated summary:** Marte et al. report that machine learning models trained on clinical and imaging data acquired within days of left-hemisphere stroke accurately forecast 12-month aphasia resolution and more modestly forecast normalization of connected-speech content. Shared and outcome-specific predictors map onto dual-stream language architecture, supporting early stratification for rehabilitation and trial enrichment.

## Introduction

Predicting which patients with acute poststroke aphasia will recover language function is central to early clinical decision-making. Recovery from acute aphasia is highly variable: approximately one-third of stroke patients present with aphasia^1^, and roughly 40% retain language deficits at 12 months^1,2^. Early and accurate prediction can guide rehabilitation intensity and timing, inform patient and family counseling, and enable prognostic enrichment of intervention trials (e.g., neuromodulation protocols^3^) with patients whose outcomes are most modifiable. The 2025 European Stroke Organisation aphasia rehabilitation guideline recommends high-dose, high-frequency speech-language therapy initiated early after stroke, reinforcing that the window for intervention-sensitive recovery may be narrow and that delays in identifying candidates for intensive treatment carry clinical cost^4^. Yet machine-learning approaches to this prediction problem have relied almost exclusively on chronic-phase features acquired months to years after stroke^5–8^, information unavailable at the acute decision point when early intensive therapy must be allocated.

The aphasia recovery literature has identified several predictive factors, among which acute severity is consistently the single most robust predictor of long-term language outcomes^9,10^ (for reviews, see^11,12^). Lazar et al. demonstrated that initial severity is a prominent predictor of chronic severity, and patients with milder initial deficits recover proportionally more completely while patients with more severe initial deficits retain larger absolute deficits at chronic timepoints even when relative improvement is substantial^9^. The RELEASE individual participant data meta-analysis^10^ confirmed that initial severity and time post-onset were the strongest predictors of recovery across language domains, explaining the majority of group-level variance in severity change, though leaving substantial individual-level variance unexplained. Wilson et al. prospectively tracked 217 patients through the first year and confirmed that acute severity is the strongest early predictor but that substantial trajectory divergence occurs between months 1 and 6 months, indicating that recovery trajectories diverge in ways not fully predicted by acute clinical presentation alone^2^. The aforementioned meta-analysis found that younger age was associated with greater recovery across language domains, whereas sex differences were not clinically meaningful and education could not be evaluated due to inconsistent reporting across datasets^10^. Other cohorts have similarly identified associations between better language outcomes and younger age^2^, higher education, and prior stroke history^11,12^. After clinical and demographic factors, lesion volume is the most consistently replicated structural predictor, correlating with both acute severity and long-term outcome but conflating damage to language-critical and non-eloquent tissue, and leaving substantial outcome variance unexplained by volume alone^13,14^. In prior cohorts, multivariable models combining acute severity, demographic factors, and lesion volume have typically explained ∼30% of variance in long-term outcomes (e.g., ^9^), leaving the majority of individual variance unexplained and motivating the integration of neuroimaging features that characterize the anatomical and network consequences of the lesion.

Neuroimaging can characterize those consequences from several perspectives. Data-driven lesion-symptom mapping across four independent cohorts has converged on a regionally specific organization of core language systems within the left perisylvian cortex: posterior superior temporal gyrus (STG) and Heschl’s gyrus damage disrupts phonological recognition and auditory comprehension; left middle and inferior temporal gyrus (MTG, ITG) damage disrupts semantic access and lexical-semantic comprehension; left supramarginal gyrus (SMG) damage disrupts phonological working memory and repetition; and left angular gyrus (AG) damage contributes to semantic integration^13^. In the frontal lobe, left pars opercularis and pars triangularis damage predicts sentence-level production and syntactic deficits^13,15^, insula and precentral damage predicts articulatory and fluency deficits^13,16^, middle frontal gyrus (MFG) damage contributes to connected-speech fluency and executive aspects of language^13^, and left superior frontal gyrus (SFG) damage shows meta-analytic evidence of implication during complex syntactic processing^17^. Using a combined longitudinal and cross-sectional design across four cohorts, Hillis et al. demonstrated that lesion load in left posterior superior temporal gyrus (pSTG) and superior longitudinal fasciculus / arcuate fasciculus (SLF/AF) predicted naming recovery through six months, independent of lesion volume^18^.

Further, white matter disconnection and network-level disruption capture how focal lesions propagate through distributed circuits^19^ (Erickson et al., 2022). The dorsal stream (SLF, AF) supports phonological-articulatory mapping, and the ventral stream (inferior fronto-occipital fasciculus [IFOF], inferior longitudinal fasciculus [ILF], uncinate fasciculus [UF]) supports semantic access, comprehension, and aspects of connected speech (Saur et al., 2008; Hickok & Poeppel, 2007). One recent connectome-based study showed that preserved efficiency of dorsal and ventral anatomical bypasses independently predicted production and comprehension, respectively^19^, converging with tract-level evidence on the importance of pathway integrity. At the network level, Hildesheim et al.^20^ used the Network Modification (NeMo) tool to retrieve “Change in Connectivity” (ChaCo) scores, a lesion-derived estimate of how much structural connectivity between brain regions is likely disrupted by superimposing the individual lesion onto a healthy-control tractographic reference set^21^, and found that connectivity disruption of left MTG and SMG predicted language production and comprehension beyond lesion volume. Interhemispheric pathways may additionally shape recovery: right-hemisphere grey matter volume in posterior temporoparietal homologues positively relates to language production (including spontaneous speech, naming, and repetition) in chronic aphasia^22^, and right-hemisphere white matter underlying the inferior frontal, precentral, and middle temporal gyri, homologues of left-hemisphere speech regions, predicts speech fluency in chronic aphasia^23^ though whether structural connectivity linking left and right homologous regions predicts differentially across longitudinal outcome dimensions has not been tested.

As noted, ML applications to aphasia prediction have been dominated by chronic-phase cohorts. Across chronic-phase studies, multimodal imaging has improved prediction over single-modality models, e.g., a multimodal model combining lesion topography, structural connectivity, and functional connectivity predicted chronic aphasia scores with cross-validated r = 0.79–0.88 across measures^5^; support vector regression (SVR) with task-based fMRI, fractional anisotropy, cerebral blood flow, and lesion load achieved r = 0.53–0.67 in 116 patients^6^; multimodal prediction of treatment response reached F1 = 0.941 with resting-state functional connectivity consistently selected among top predictors^7^; and nested-cross validation (CV) SVR with resting-state fMRI and structural integrity achieved r = 0.70 in 76 patients^8^. However, not all studies agree on the incremental value of connectivity features over lesion data alone^24^, and all of these studies relied on information unavailable at the acute decision point: network reorganization, functional compensation, and bilateral recruitment that develop over weeks to months after stroke. Levy et al.^25^ addressed this gap in part, using SVR-based multivariate lesion-symptom mapping in 217 patients; their models predicted ∼60% of one-year aphasia severity variance from acute lesion location and demographics, with initial severity adding further incremental variance beyond these structural and demographic features.

Finally, nearly all the ML prediction studies reviewed above predict a single composite outcome, yet communication in daily life (i.e., conversations, narrating events, explaining symptoms to a physician) requires connected speech that depends on simultaneous lexical retrieval, syntactic assembly, and discourse-level planning. Content unit analysis partially captures this dimension by indexing propositional content in naturalistic production. Bunker et al.^26^ demonstrated that patients above the WAB-AQ diagnostic cutoff still showed reduced content production relative to healthy controls on the Modern Cookie Theft picture description^27^, confirming that meaningful residual impairments persist in patients classified as having “resolved” aphasia by standard criteria. Other studies have reinforced the sensitivity of content-based measures to these residual deficits, demonstrating that main concept production distinguishes “subclinical” from non-clinical speakers across multiple discourse tasks in a large normative sample^28^. Critically, subacute aphasia profiles have been shown to resolve into three orthogonal components – fluency, semantic/executive, and phonology – whose longitudinal recovery trajectories between 2 weeks and 4 months post-stroke are uncorrelated^29^; the fluency component (i.e., the component most relevant to connected speech production) followed a trajectory distinct from the others, implying that models optimized for a single severity composite may systematically miss the component where recovery is most constrained. Neuroimaging evidence further suggests that the neural substrates of discourse may differ in emphasis from those of global severity, such that impaired discourse content has been linked specifically to frontal white matter damage in chronic aphasia^30^, and a lexical-semantic discourse factor has been mapped onto a left inferior parietal, superior/middle temporal, and lateral occipital cluster in voxel wise lesion-symptom mapping^31^. Whether the predictors of discourse normalization differ from those of global severity resolution, and at which level of the imaging hierarchy the differences emerge, has not been tested in an acute-phase predictive framework.

To address these gaps and examine the feasibility of early prediction from acute-phase data, we used ML to predict two 12-month outcomes from acute-phase features in 73 patients with left-hemisphere stroke and aphasia: aphasia resolution (Western Aphasia Battery–Revised Aphasia Quotient [WAB-AQ] ≥ 93.8) and discourse content normalization (Modern Cookie Theft picture description content units [CUs] ≥ 22.1; Berube et al., 2019). A hierarchical feature set design (clinical–volume–anatomy–network) tests the contribution of each modality incrementally. We compared four algorithms (Ridge, SVR, random forests [RF], eXtreme Gradient Boosting [XGBoost]) to assess whether model flexibility (i.e., the capacity to capture nonlinear severity-outcome relationships) influences prediction beyond feature composition. We used nested cross-validation and SHapley Additive exPlanations (SHAP) stability analysis for model interpretation^8,32^. We hypothesized that (1) acute-phase data would predict both aphasia resolution with high accuracy and discourse normalization more modestly, driven primarily by acute WAB-AQ, consistent with the established predictive power of initial severity^9,10^; (2) clinical features alone would achieve strong prediction for aphasia resolution, with imaging adding incrementally, given the tight coupling between severity and lesion extent at the acute timepoint; (3) SHAP analysis would reveal common predictors (acute severity, lesion volume) alongside outcome-specific features, with ventral stream tracts (IFOF, ILF) prominent for WAB-AQ resolution (which weights comprehension and naming) and a more distributed profile including dorsal stream, mapping the convergence and divergence of predictors onto dual-stream architecture^33^.

## Materials and methods

### Participants

Participants were 73 patients with acute left-hemisphere (LH) ischemic stroke and aphasia, drawn from a longitudinal program at Johns Hopkins Hospital that enrolled 617 English-speaking adults with acute LH ischemic stroke between 2007 and 2024. Inclusion criteria for the present analysis were confirmed LH ischemic stroke on magnetic resonance imaging (MRI), acute WAB-AQ assessment within the first week post-onset (mean 2.8 days, SD = 2.2), completion of 12-month follow-up, and a complete neuroimaging processing pipeline. Patients were excluded from longitudinal monitoring if they had strokes limited to the brainstem or cerebellum, hemorrhagic stroke, preexisting neurological diseases that affect language and cognition (e.g., dementia), intellectual disability, or severe or uncorrected vision or hearing loss. Because the core language battery and discourse protocol were updated across this enrollment window (the WAB-R was adopted as the standard aphasia assessment following earlier diagnostic protocols, and the picture-description task used to derive discourse measures was introduced at a later phase of the program), paired acute and 12-month WAB-AQ were available for an analytic subset of the full enrollment, and discourse measures for a smaller subset still; all analyses use the harmonized measures available across phases. Demographic and clinical characteristics and between-group comparisons are reported in Table 1; lesion distribution is shown in Figure 1A. At 12 months, 47 of 73 patients (64.4%) met aphasia resolution criteria, and 28 of 61 (45.9%) met discourse normalization criteria. All study procedures were approved by the Johns Hopkins University School of Medicine institutional review board (protocol NA_00042097), and informed written consent was obtained by participants or their legally authorized representatives as appropriate prior to participation.

**Figure 1A-C.**
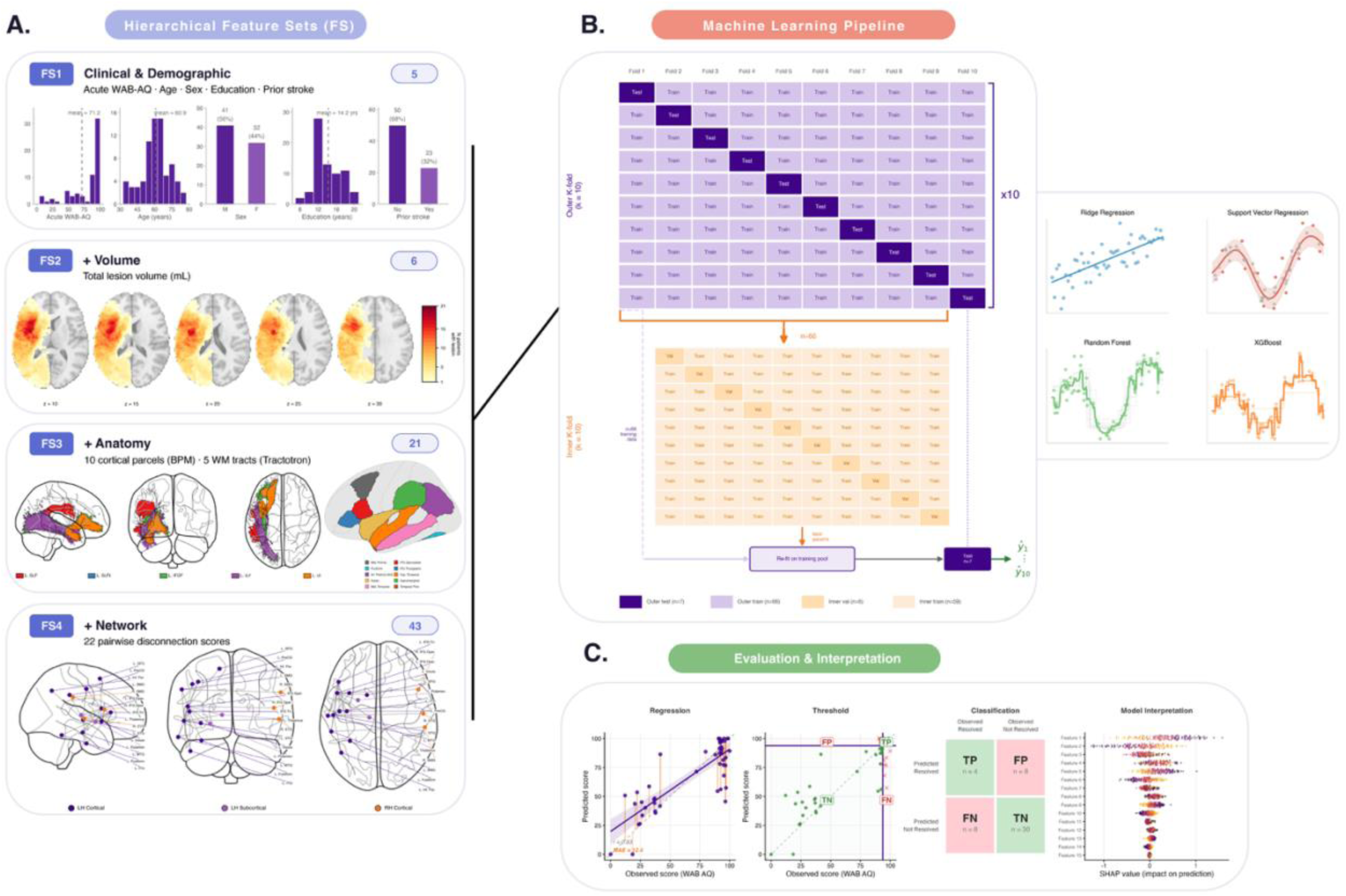
Analytic pipeline overview. (A) Hierarchical feature sets FS1-FS4, with cumulative feature counts shown in each panel’s upper-right badge. FS1 (5 features): acute WAB-AQ, age, sex, education, and prior stroke, with sample distributions. FS2 (6 features) adds total lesion volume, illustrated by the lesion overlap map across 73 patients on axial MNI152 slices. FS3 (21 features) adds 10 LH BPM cortical parcels and 5 Tractotron tracts (SLF, arcuate fasciculus, IFOF, ILF, UF), shown on cortical surface renderings. FS4 (43 features) adds 22 pairwise NeMo language-circuit disconnection scores on the fs86 parcellation, depicted as connectivity pairs spanning LH cortical, LH subcortical, and RH cortical nodes. (B) Nested cross-validation structure. The outer loop runs 10-fold CV repeated 10 times (100 outer test folds total); each outer fold holds out ∼7 patients for testing (purple) and trains on the remainder (light purple). Within each outer training fold, an inner 10-fold grid-search CV (orange) tunes hyperparameters, partitioning the outer training data into inner training (light orange) and inner validation (orange) sets. Best hyperparameters are refit on the full outer training pool and evaluated on the held-out outer test fold. Predictions from all outer test folds across 10 repeats (ŷ_1_…ŷ_10_) are aggregated. Four algorithms are evaluated: Ridge regression, SVR, RF, and XGBoost. (C) Evaluation framework (illustrative values shown). Continuous regression predictions (left; predicted vs. observed scores with Pearson r, R², and MAE) are converted to binary classifications by applying the clinical threshold, yielding the confusion matrix of true positives (TP), false positives (FP), false negatives (FN), and true negatives (TN). Interpretation is performed via SHAP analysis per feature across cross-validation folds (right). *Alt text: Three-panel schematic of the analytic pipeline. Panel A shows four hierarchical feature sets building from clinical-only (FS1, five features) to clinical-plus-network (FS4, 43 features), with cumulative feature count shown in each panel’s upper-right badge and illustrative data visualizations (demographic distributions, lesion overlap map, cortical parcels and tracts, connectivity matrix). Panel B illustrates the nested cross-validation structure, with an outer 10-fold loop repeated 10 times for performance evaluation and an inner 10-fold grid-search loop for hyperparameter tuning. Panel C depicts the four machine-learning algorithms compared (Ridge regression, support vector regression, random forest, XGBoost) and the SHAP-based feature-importance output*.

**Table 1.** Baseline Characteristics by 12-month Aphasia Status.

| Characteristic | Overall<br>(n=73) | Resolved<br>(n=47) | Not Resolved<br>(n=26) | p |
| --- | --- | --- | --- | --- |
| Age, y | 60.9 (11.9) | 59.8 (12.5) | 63.0 (10.5) | 0.3 |
| Sex, % female | 43.8 | 36.2 | 57.7 | 0.090 |
| Race, % |  |  |  | 0.4 |
| Asian | 1.4 | 2.1 | 0.0 |  |
| Black | 58.9 | 55.3 | 65.4 |  |
| Other | 1.4 | 0.0 | 3.8 |  |
| White | 38.4 | 42.6 | 30.8 |  |
| Education, y | 14.3 (2.9) | 14.8 (3.0) | 13.3 (2.4) | 0.033 |
| Prior stroke, % | 31.5 | 31.9 | 30.8 | >0.9 |
| Acute WAB-AQ | 71.3 (34.5) | 89.8 (18.1) | 37.8 (31.8) | <0.001 |
| Lesion volume, mL | 36.7 (65.9) | 13.1 (22.4) | 79.2 (93.1) | 0.001 |
| Days post onset | 2.8 (2.2) | 2.8 (2.0) | 2.9 (2.7) | >0.9 |

Note: Mean (SD) or %. p values from Welch two-sample t-test (continuous) or Fisher’s exact test (categorical).

### Outcome Measures

Two binary outcomes were defined a priori from continuous 12-month scores. Aphasia resolution was defined as WAB-AQ ≥ 93.8^34^; the WAB-R is a standardized aphasia assessment in which the AQ (0–100) summarizes performance across spontaneous speech (a composite of fluency and information content scales), auditory comprehension, repetition, and naming subtests, with AQ ≥ 93.8 the standard cutoff for clinically meaningful resolution of aphasia. Discourse normalization was defined as Modern Cookie Theft picture description content units (CUs) ≥ 22.1. The Modern Cookie Theft is a quantified picture-description task based on an updated version of the Boston Diagnostic Aphasia Examination Cookie Theft picture depicting a kitchen scene with children, caregiver, and dog^27^. Content units are concepts mentioned by at least three of 50 neurologically healthy controls in describing the picture; the 22.1 cutoff corresponds to approximately one standard deviation below the healthy control mean (mean 33.5 ± 11.4 CUs)^27^. The two outcomes capture complementary dimensions of language: a global severity composite and a naturalistic production measure.

### Neuroimaging Processing

Baseline imaging was acquired at the time of admission for the index stroke on a 3 Tesla scanner; all patients underwent diffusion-weighted imaging, from which ischemic lesion volume was derived. Acute diffusion-weighted MRI sequences were processed with the Acute-stroke Detection Segmentation (ADS) pipeline^35^, which generated an initial lesion mask via deep-learning segmentation on diffusion-weighted imaging. Masks were reviewed and manually corrected in ITK-SNAP^36^ by a trained research assistant (M.C.) blinded to 12-month outcome and subsequently verified by the lead author (M.J.M.). From each lesion mask, we derived three imaging feature types at successive levels of granularity. First, proportional damage to each cortical parcel of the Brain Parcellation Map (BPM)^37^ was computed via ADS. Second, disconnection probability (0–1) for each tract in the JHU white matter atlas^38^ was estimated with Tractotron^39^ from the spatial overlap between the lesion and normative tract probability maps. Third, pairwise ChaCo scores on the fs86 Desikan–Killiany parcellation^40^ were computed with the NeMo tool^21^, estimating connectivity change between region pairs attributable to the lesion. All BPM, Tractotron, and NeMo features were residualized against lesion volume within each cross-validation fold so that any contribution of a specific parcel, tract, or disconnection pair reflects signal beyond total lesion extent^41^.

### Feature Sets

Four hierarchical feature sets (FS1–FS4) were defined *a priori* to isolate the incremental predictive contribution of each imaging modality beyond clinical severity. Here, feature counts are cumulative. FS1 (clinical, 5 features): acute WAB-AQ, age, sex, education, and prior stroke. FS2 adds lesion volume (6 features total). FS3 adds 10 LH cortical parcels and 5 LH white matter tracts selected from the perisylvian network and dual-stream model^33^ (21 features total). Cortical parcels span the perisylvian network (pars opercularis, pars triangularis, STG, SMG, angular gyrus, middle temporal gyrus, inferior temporal gyrus, insula, superior frontal gyrus, middle frontal gyrus); tracts span the dorsal stream (SLF, arcuate fasciculus) and ventral stream (IFOF, ILF, uncinate fasciculus). FS4 adds 22 pairwise NeMo language-circuit disconnection scores spanning dorsal, receptive, production, interhemispheric, and extended semantic networks (43 features total; Supplementary Table S3). The intention was to test whether each modality contributes predictive variance beyond the preceding layers while preserving all prior features. Region-, tract-, and circuit-selection are additionally detailed in Supplementary S1, S2, and Section S3, respectively.

### Machine Learning Pipeline

The nested cross-validation structure follows current best-practice recommendations for neuroimaging prediction^42,43^. Four algorithms were tested: Ridge regression, SVR with a radial basis function (RBF) kernel, RF, and XGBoost (see Supplementary S4 for selection rationale). All four were implemented as regressors predicting continuous 12-month outcomes, with clinical thresholds applied post-hoc to derive binary classifications. A regression-then-threshold approach uses the full gradient of recovery during training rather than discarding it at an arbitrary cut-point and recovers the clinically meaningful binary decision only at the point of interpretation^44,45^.

All preprocessing was performed inside each training fold to prevent leakage: z-score standardization of features, residualization of imaging features against lesion volume, and inverse-density sample weighting. Twelve-month outcome distributions for both WAB-AQ and content units were left-skewed, i.e., most patients approached ceiling, whereas the tail of poor-outcome cases was sparse, so that standard loss functions would let models optimize the majority at the expense of the clinically most-relevant subgroup. To counter this, inverse-density weighting was applied using Gaussian kernel density estimation (KDE) of the outcome distribution, with weights mean-normalized and capped at 5× the median to prevent individual-case dominance (see Supplementary S5 for full parameters).

The nested cross-validation structure (see Figure 1B) ensures that no test-fold information influences feature selection, hyperparameter tuning, or preprocessing parameters, and used 10-fold cross-validation repeated 10 times (100 outer test folds total) for the outer evaluation loop and 10-fold grid-search cross-validation for the inner hyperparameter optimization loop (Supplementary Table S1). Outer folds were stratified on outcome quartiles to preserve the distribution of severe cases across folds^43^. Recursive feature elimination (RFE; step = 0.1) was applied within-fold to FS3 and FS4 to reduce dimensionality while retaining predictive signal^5^.

All analyses were conducted in Python 3.11 (scikit-learn 1.4, XGBoost 2.0, SHAP 0.44, NumPy 1.26, SciPy 1.11). Code to reproduce all analyses is available at https://osf.io/m8uv2/overview?view_only=0e0f6c5e75cd4f198c50fa23c3e34a16.

### Model Evaluation

Models were evaluated on continuous fit (Pearson r, R², mean absolute error [MAE]) and binary classification (F1, sensitivity, specificity, positive predictive value [PPV], negative predictive value [NPV], balanced accuracy, Matthews Correlation Coefficient [MCC]; for definitions and rationale, see Supplementary S6). F1 was the primary selection metric because it penalizes models that achieve one of PPV or sensitivity at the expense of the other; MCC and balanced accuracy are reported because both adjust for the 64.4% resolution base rate^46^. 95% bootstrap CIs were computed from 2,000 resamples (percentile method), and permutation-based p-values from 1,000 label-shuffled pipeline re-runs. The complete 32-model outcomes (4 algorithms × 4 feature sets × 2 outcomes) are reported in Supplementary Table S2. Reporting follows the TRIPOD+AI guideline (see Supplementary S6). Within-subject SD of predicted scores across the 10 cross-validation repeats was compared between correctly and incorrectly classified patients with two-sided Wilcoxon rank-sum tests.

#### Model Interpretation

SHAP values^32^ were computed for the top-performing model per outcome. Because a feature’s importance in any single cross-validation fold can be driven by the particular training split, we ranked features by their mean |SHAP| across all 100 outer test folds (10 folds × 10 repeats), such that features that consistently rank highly across independent partitions are less likely to reflect idiosyncratic fold-level noise. Full computation and stability protocol are detailed in Supplementary S7.

## Results

### Aphasia Resolution

The FS4 RF model achieved the highest F1 across all feature sets for aphasia resolution (n = 73; 47 resolved, 26 not resolved) with F1 = 0.874 (95% CI, 0.800–0.941), a sensitivity of 0.809 (95% CI, 0.700–0.913), i.e., the model correctly identified 81% of patients who went on to resolve, a specificity of 0.923 (95% CI, 0.818–1.000), i.e., among patients who did not resolve, 92% were correctly flagged as non-resolvers, and PPV of 0.950 (95% CI, 0.878–1.000). The confusion matrix showed 38 true positives, 24 true negatives, 2 false positives, and 9 false negatives; of 73 patients the model correctly triaged 62 (85%). NPV was 0.727 (95% CI, 0.581–0.861). Balanced accuracy was 0.866 (95% CI, 0.787–0.938) and MCC was 0.704 (95% CI, 0.548–0.842), both exceeding the 0.500 and 0.000 expected from a naive classifier at the 64.4% base rate (permutation p < .001). The FS4 RF model predicted continuous WAB-AQ with r = 0.827 (95% CI, 0.678–0.937), MAE = 7.26 (95% CI, 4.56–10.06) points, and R² = 0.652 (95% CI, 0.154–0.873; Figures 2A and 3A); within-subject SD of predictions did not differ between correctly and incorrectly classified patients (Figure 2B; Wilcoxon rank-sum p = .118). Performance was stable across feature sets, with RF F1 ranging from 0.851 to 0.874 from FS1 to FS4 (for full model performance values, see Supplementary Table S2).

**Figure 2A-D.**
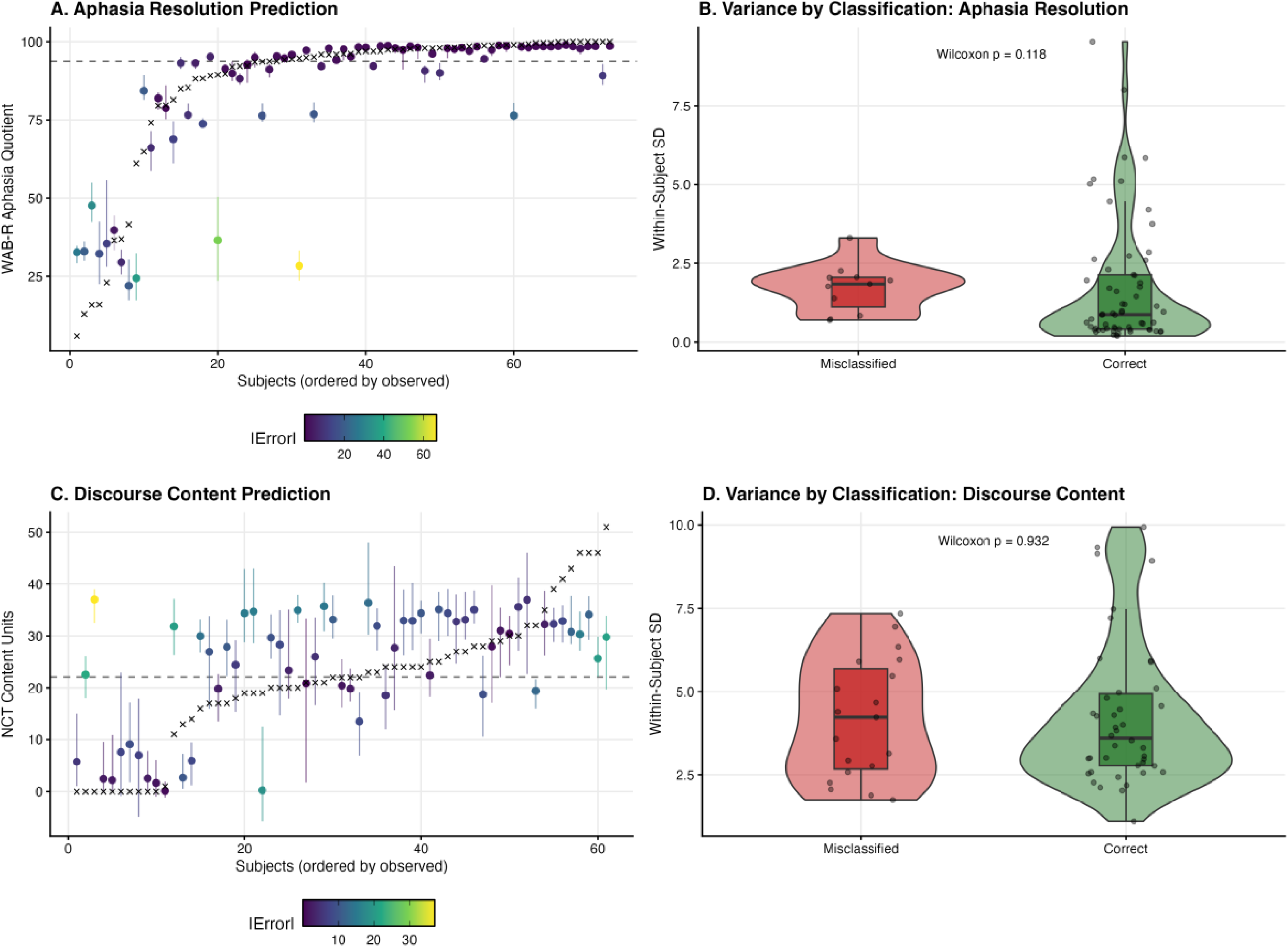
Model performance and prediction stability. (A) Aphasia resolution (FS4 RF, n = 73): per-subject predicted versus observed 12-month WAB-AQ, ordered along the x-axis by observed score. Filled circles show the mean cross-fold prediction per subject; vertical bars show the within-subject prediction range across 10 cross-validation repeats; × markers show observed scores. Points are colored by absolute prediction error. (B) Aphasia resolution: within-subject SD of predicted scores, partitioned by classification outcome (correctly classified in green, misclassified in red). (C) Discourse content normalization (FS4 SVR, n = 61): per-subject predicted versus observed 12-month content units, plotted as in (A). (D) Discourse content normalization: within-subject SD of predicted scores, plotted and tested as in (B). *Alt text: Four-panel summary of model performance and prediction stability. Panels A and C plot per-subject predictions against observed scores for aphasia resolution (A, n = 73) and discourse content normalization (C, n = 61), ordered along the x-axis by observed score, with filled circles showing mean cross-fold predictions, vertical bars the within-subject prediction range across cross-validation repeats, and × markers the observed scores, colored by absolute prediction error. Panels B and D show within-subject prediction standard deviations partitioned by classification outcome, with correctly classified subjects in green and misclassified subjects in red*.

**Figure 3A-B.**
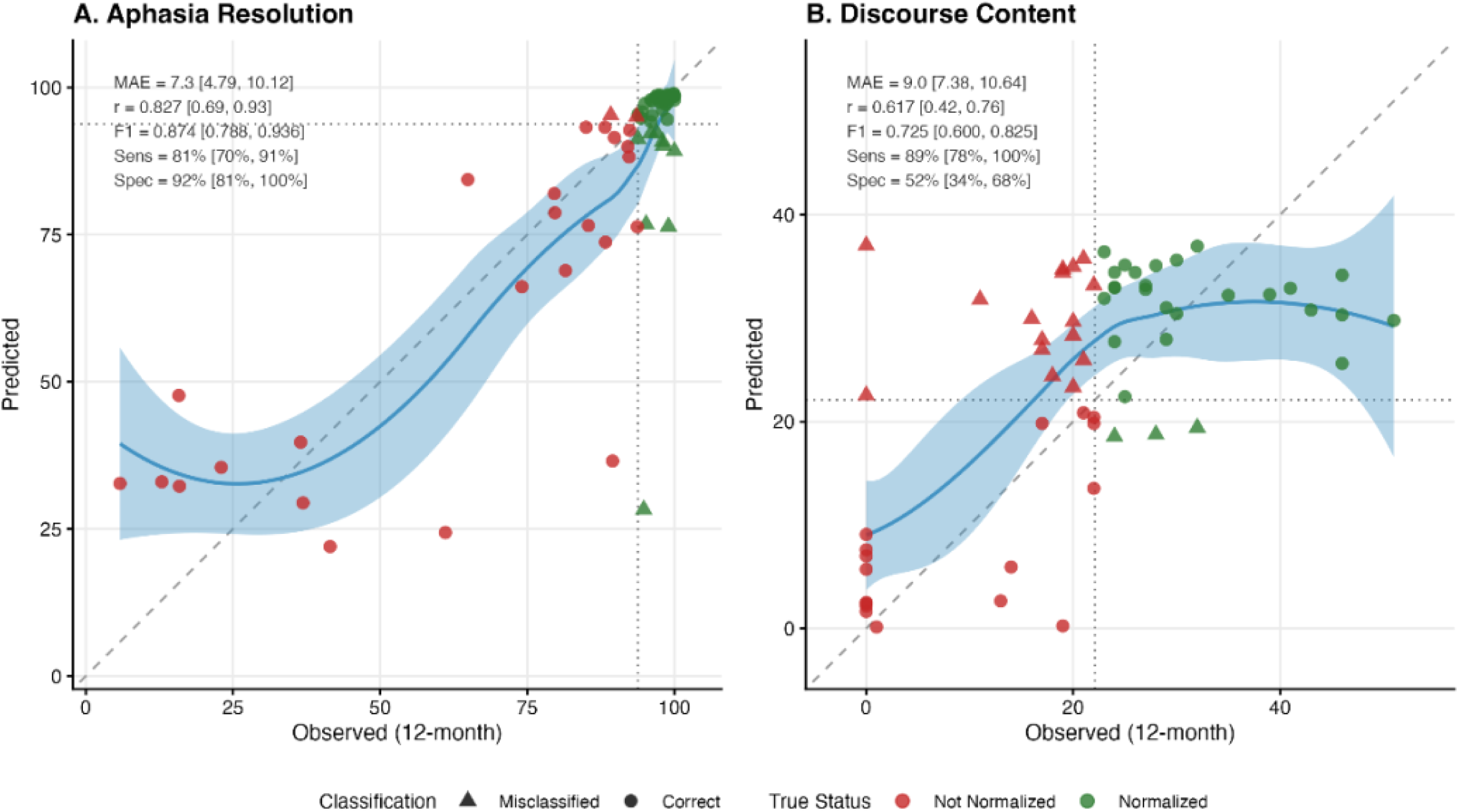
Predicted versus observed 12-month scores for the top-performing models. (A) Aphasia resolution (FS4 RF, n = 73): continuous WAB-AQ predictions against observed 12-month WAB-AQ. (B) Discourse content normalization (FS4 SVR, n = 61): continuous content unit (CU) predictions against observed 12-month CUs. Point shape encodes binary classification under the clinical threshold (circles = correctly classified; triangles = misclassified) and color encodes true recovery status (green = resolved/normalized; red = not resolved/not normalized). Dashed lines show clinical thresholds; solid diagonal shows perfect prediction. *Alt text: Two-panel scatter plot of continuous predicted versus observed 12-month scores for the top-performing models. Panel A plots aphasia resolution predictions against observed Western Aphasia Battery-Revised Aphasia Quotient scores (n = 73). Panel B plots discourse predictions against observed. Modern Cookie Theft content-unit counts (n = 61). Point shape encodes binary classification under the clinical threshold (circles = correctly classified; triangles = misclassified); color encodes true recovery status (green = resolved or normalized; red = not resolved or not normalized). Dashed lines mark the clinical thresholds; solid diagonals mark perfect prediction.*

### Aphasia Resolution Feature Importance

For aphasia resolution, acute WAB-AQ was the top-ranked feature with mean |SHAP| = 13.60, approximately 20× the next feature (Table 3; Supplementary Figure S1A). Three features appeared in the top 10 for both outcomes: acute WAB-AQ, lesion volume, and left pars triangularis (Table 3). Lesion volume was the second-ranked feature for aphasia resolution (0.687). The top-ranked cortical parcels were pars triangularis (0.255), middle frontal gyrus (0.203), insula (0.179), and pars opercularis (0.146), and the top-ranked white matter tracts were IFOF (0.249) and ILF (0.208). Among network features, frontal-circuit connectivity was most important, i.e., intrafrontal L.POP–L.PTR (0.233) and thalamofrontal L.thalamus–L.POP (0.198). See Figure 4A-C for visualization; the complete stability-ranked top 15 features and SHAP bar summary are shown in Supplementary Figure S1A.

**Figure 4A-F.**
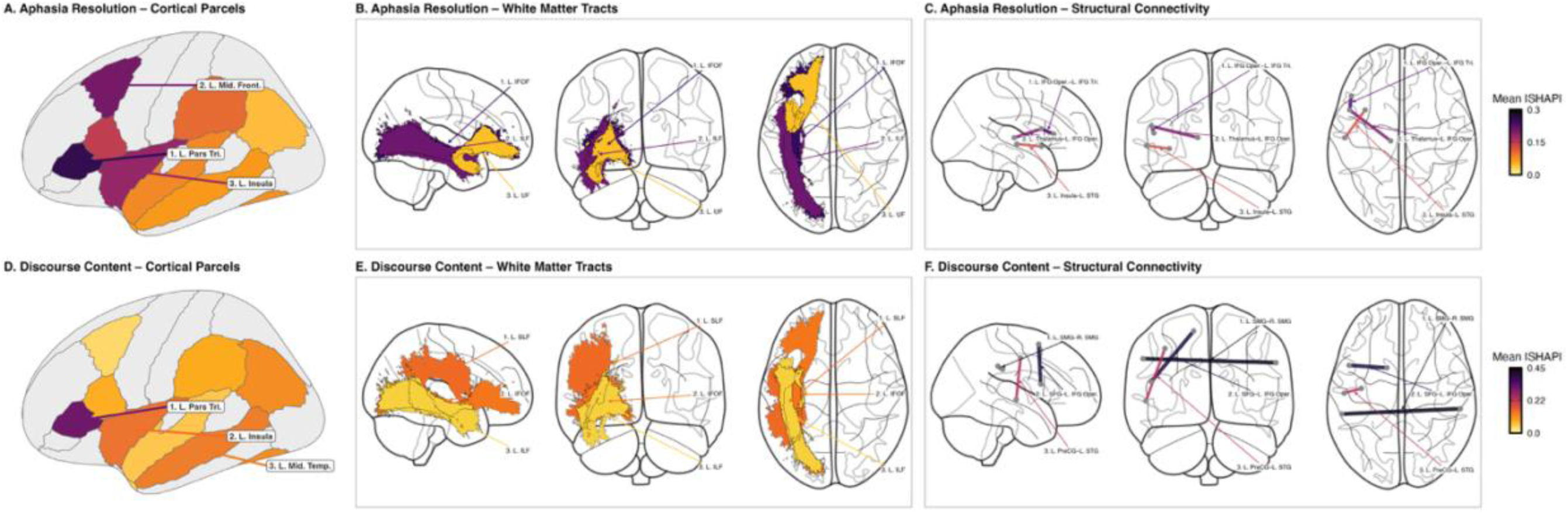
Top SHAP-ranked imaging features for each outcome visualized on anatomical templates. For each outcome and each imaging class, the three features with the highest mean |SHAP| across all 100 outer test folds are shown, shaded by their mean |SHAP| value. (A–C) Aphasia resolution (FS4 RF): (A) left-hemisphere cortical parcels; (B) left-hemisphere white matter tracts (top three: IFOF, ILF, UF); (C) structural connectivity pairs (top three: pars opercularis–pars triangularis, L.thalamus–pars opercularis, insula–STG). (D–F) Discourse content normalization (FS4 SVR): (D) left-hemisphere cortical parcels; (E) left-hemisphere white matter tracts (top three: SLF, IFOF, ILF); (F) structural connectivity pairs including right-hemisphere homologues (top three: L.SMG–R.SMG, SFG–pars opercularis, precentral–STG). *Alt text: Six-panel anatomical visualization of the top-ranked imaging features for each outcome, with feature shading scaled by mean absolute SHAP value. Panels A-C show features for aphasia resolution: left-hemisphere cortical parcels (A), left-hemisphere white-matter tracts (B), and structural connectivity pairs (C). Panels D-F show the same three feature classes for discourse content normalization*.

### Discourse Content Normalization

For discourse content normalization (n = 61; 28 normalized, 33 not), the FS4 SVR model achieved the highest F1 at 0.725 (95% CI, 0.593–0.831), with sensitivity of 0.893 (95% CI, 0.767–1.000), i.e., the model identified 89% of patients who went on to normalize, and specificity of 0.515 (95% CI, 0.343–0.684), i.e., among patients who did not normalize, roughly half were correctly flagged (Table 2). The confusion matrix showed 25 true positives, 17 true negatives, 16 false positives, and 3 false negatives; of 61 patients the model correctly triaged 42 (69%). Balanced accuracy was 0.704 (95% CI, 0.602–0.800) and MCC was 0.433 (95% CI, 0.220–0.647; permutation p = .003). The model predicted CUs with r = 0.617 (95% CI, 0.437–0.753), MAE = 8.96 (95% CI, 7.39–10.64) CUs, and R² = 0.252 (95% CI, -0.164–0.512; Figures 2C and 3B); within-subject SD of predictions did not differ between correctly and incorrectly classified patients (Figure 2D; Wilcoxon rank-sum p = .932). Performance was stable across feature sets, with a modest ΔF1 = +0.029 from FS1 to FS4 (Supplementary Table S2).

**Table 2.**
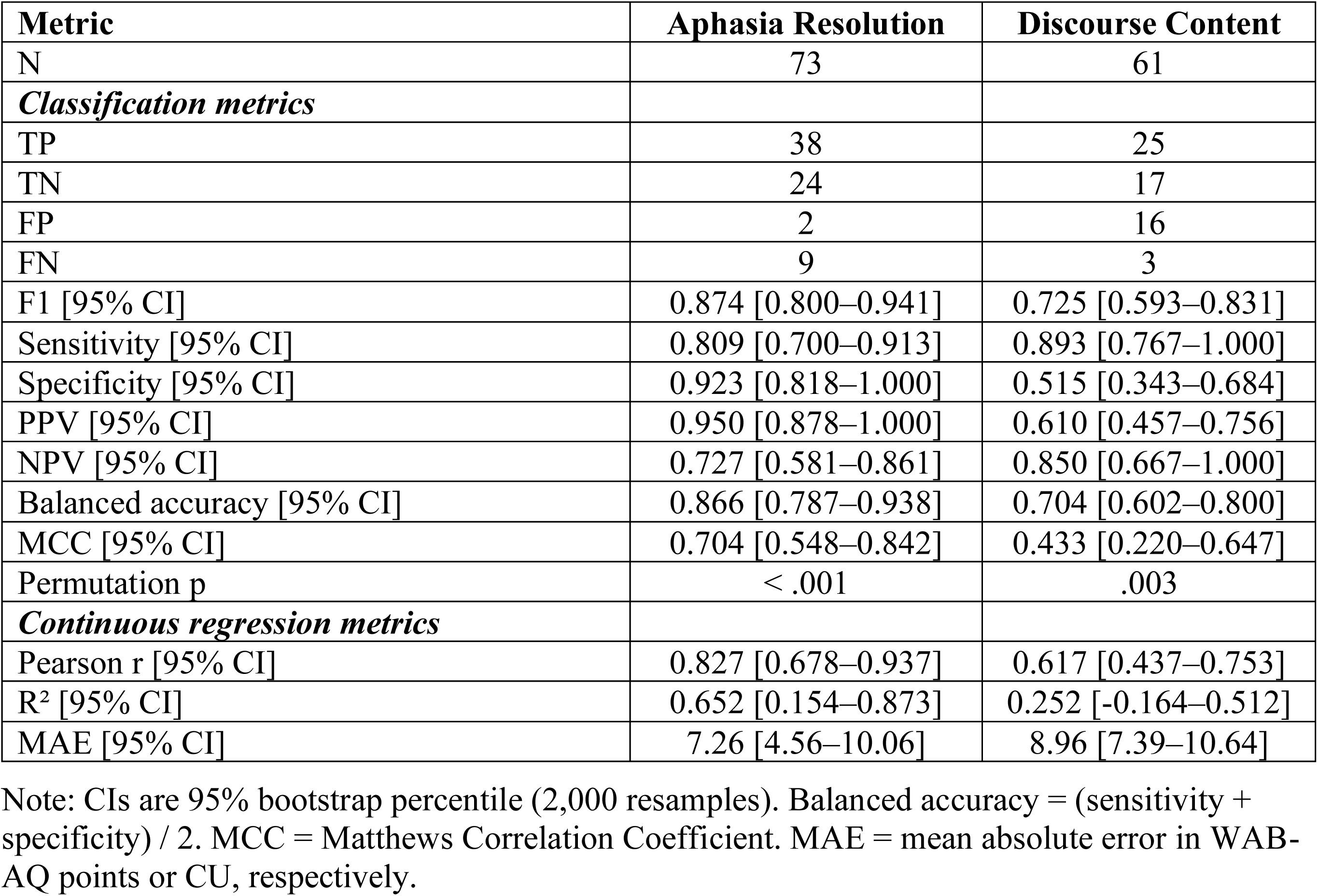
Model Performance.

| **Metric** | **Aphasia Resolution** | **Discourse Content** |
| --- | --- | --- |
| N | 73 | 61 |
| ***Classification metrics*** |  |  |
| TP | 38 | 25 |
| TN | 24 | 17 |
| FP | 2 | 16 |
| FN | 9 | 3 |
| F1 [95% CI] | 0.874 [0.800–0.941] | 0.725 [0.593–0.831] |
| Sensitivity [95% CI] | 0.809 [0.700–0.913] | 0.893 [0.767–1.000] |
| Specificity [95% CI] | 0.923 [0.818–1.000] | 0.515 [0.343–0.684] |
| PPV [95% CI] | 0.950 [0.878–1.000] | 0.610 [0.457–0.756] |
| NPV [95% CI] | 0.727 [0.581–0.861] | 0.850 [0.667–1.000] |
| Balanced accuracy [95% CI] | 0.866 [0.787–0.938] | 0.704 [0.602–0.800] |
| MCC [95% CI] | 0.704 [0.548–0.842] | 0.433 [0.220–0.647] |
| Permutation p | < .001 | .003 |
| ***Continuous regression metrics*** |  |  |
| Pearson r [95% CI] | 0.827 [0.678–0.937] | 0.617 [0.437–0.753] |
| R<sup>2</sup> [95% CI] | 0.652 [0.154–0.873] | 0.252 [-0.164–0.512] |
| MAE [95% CI] | 7.26 [4.56–10.06] | 8.96 [7.39–10.64] |

Note: CIs are 95% bootstrap percentile (2,000 resamples). Balanced accuracy = (sensitivity + specificity) / 2. MCC = Matthews Correlation Coefficient. MAE = mean absolute error in WAB-AQ points or CU, respectively.

**Table 3.** SHAP Feature Importance Summary.

| Rank | WAB Feature | \|SHAP\| | Modality | CU Feature | \|SHAP\| | Modality |
| --- | --- | --- | --- | --- | --- | --- |
| 1 | Acute WAB-AQ | 13.60 | Clinical | Acute WAB-AQ | 1.96 | Clinical |
| 2 | Lesion volume | 0.687 | Volume | Lesion volume | 0.850 | Volume |
| 3 | L pars triangularis | 0.255 | Cortical | Prior stroke | 0.595 | Clinical |
| 4 | IFOF | 0.249 | WM | L.SMG–R.SMG | 0.432 | Network |
| 5 | L.POP–L.PTR | 0.233 | Network | Sex | 0.431 | Clinical |
| 6 | ILF | 0.208 | WM | L.SFG–L.POP | 0.401 | Network |
| 7 | L MFG | 0.203 | Cortical | Age | 0.392 | Clinical |
| 8 | L.thalamus–<br>L.POP | 0.198 | Network | Education | 0.345 | Clinical |
| 9 | L insula | 0.179 | Cortical | L pars triangularis | 0.313 | Cortical |
| 10 | L pars opercularis | 0.146 | Cortical | L.PrCG–L.STG | 0.234 | Network |

Note: Values are mean absolute SHAP averaged across 100 outer test folds (10 folds × 10 repeats). Modality: Clinical (acute severity + demographics), Volume (lesion volume, mL), Cortical (BPM parcel), WM (Tractotron tract), Network (NeMo connection pair). WAB = Western Aphasia Battery–Revised Aphasia Quotient; CU = content units.

### Discourse Content Feature Importance

For discourse normalization, the SHAP ranking was more distributed across feature classes (Figure 4D–F; Supplementary Figure S1B). Demographic (FS1) features rose relative to aphasia resolution, i.e., prior stroke (0.595), sex (0.431), age (0.392), and education (0.345) were all in the top 10. Beyond lesion volume (FS2), left pars triangularis was the only FS3 cortical parcel to reach the top 10 (0.313), and no individual tract did so. Network features were however present, led by interhemispheric SMG connectivity (L.SMG–R.SMG, 0.432), L.SFG–L.POP (0.401), and L.PrCG–L.STG (0.234). Among tracts, SLF (0.148), IFOF (0.137), and ILF (0.050) were the three top-ranked (see Figure 4D-E).

## Discussion

Three findings ground this work and jointly address our aims. First, acute-phase features predicted 12-month aphasia resolution with high accuracy (F1 = 0.874; MAE = 7.26 WAB-AQ points), confirming that information available at bedside within days of stroke is sufficient for an actionable one-year forecast given comprehensive acute language assessment. Second, clinical features alone reached near-ceiling performance (F1 = 0.851), with imaging adding only small aggregate gains (ΔF1 = +0.023 for aphasia resolution; +0.029 for discourse) that nonetheless carried interpretive value. Third, SHAP rankings revealed a tripartite architecture: acute WAB-AQ, lesion volume, and damage to left pars triangularis beyond what lesion volume alone captured underlay both outcomes, while further imaging predictors diverged. Ventral-stream white matter and left inferior frontal circuitry was linked to aphasia resolution, whereas discourse drew additionally on dorsal-stream, interhemispheric, and prefrontal predictors. The two outcomes therefore tap partially distinct substrates rather than differing only in severity.

The shared predictors recapitulate the most robust findings in the prior literature, namely initial severity, lesion volume, and left inferior frontal damage^9,10,13,18^. That these rose to the top for both outcomes in a data-driven ranking, not only for the composite WAB-AQ, indicates that they index a general severity-gradient substrate rather than an outcome-specific one.

Beyond this shared core, aphasia resolution was predicted by ventral-stream white matter (IFOF, ILF) and a left inferior frontal production circuit (pars triangularis, pars opercularis, their intrafrontal connectivity, and thalamocortical coupling to pars opercularis). This pattern is consistent with the composition of the WAB-AQ, which weights comprehension and naming (operations that draw on ventral-stream semantic access) and with connectome-bypass evidence that preserved ventral-stream efficiency predicts comprehension while dorsal-stream efficiency predicts lexical production^13,15,19^. Thalamocortical coupling to pars opercularis has a direct empirical counterpart in recent lesion-network mapping, which implicates the left ventrolateral and ventral anterior thalamic nuclei in engaging Broca’s area during language processing^47^. Preservation of this coupling at the acute phase may therefore support recovery of the lexical-retrieval operations that the WAB-AQ indexes.

Discourse normalization was predicted with more modest accuracy, so the architecture that follows bears more speculatively on mechanism than the aphasia-resolution profile. That said, discourse predictors were markedly more distributed. Demographic features (prior stroke, sex, age, education) rose into the top 10, though most the distinctive discourse-specific set of features were interhemispheric L.SMG–R.SMG connectivity, with SFG–pars opercularis and precentral–STG also rising. Three converging lines of evidence support a structural-substrate account of this pattern. Right temporoparietal grey matter is positively associated with speech production, but not comprehension, in chronic aphasia, with evidence of post-stroke hypertrophy^22^. Right-hemisphere white matter integrity in inferior frontal, precentral, and middle temporal homologues predicts chronic speech fluency^23^. Further, subacute fluency-component recovery tracks increasing activation in right temporo-occipital MTG and bilateral MFG^29^. The prefrontal contribution converges with evidence that impaired discourse content in chronic aphasia localizes to frontal white matter damage^30^, and the inferior-parietal contribution accords with recent lesion-symptom mapping of a lexical-semantic discourse factor onto left inferior parietal and superior/middle temporal cortex^31^. Acute interhemispheric temporoparietal connectivity may thus index a structural precondition for later right-hemisphere recruitment, a process that is regionally heterogeneous rather than uniform across homotopic nodes, with recruitment constrained by both lesion location and preserved connectivity^48^. A falsifiable prediction follows, e.g., baseline L.SMG–R.SMG connectivity should interact with subsequent right-SMG fMRI recruitment in accounting for discourse recovery, distinguishing structural-substrate accounts from purely functional-recruitment accounts.

Next, a broader implication emerges from the comparability of these acute-phase models to chronic-phase multimodal ones (c.f., ^5–8^), in that the structural substrate present within days of stroke may constrain the functional reorganization that unfolds over weeks to months. This reading is consistent with longitudinal evidence that acute white matter integrity predicts later connected-speech recovery^49^, that regions with stronger baseline lesion-connectivity show steeper subacute language reactivation^50^, and that post-stroke language reorganization predominantly occurs within pre-existing networks rather than through novel recruitment^51^. If the acute network sets much of the recovery trajectory, the marginal prognostic yield of later multimodal imaging may be narrower than assumed.

It is also notable that algorithm choice mattered more than feature composition for aphasia resolution. Tree-based nonlinear models (RF, XGB) outperformed linear models on the same five clinical features, reflecting a threshold-like severity–outcome surface in which mild deficits almost always resolve, severe deficits rarely do, and the most predictive uncertainty lives in the midrange where nonlinearity matters. A larger acute cohort (n = 217) reached comparable variance explained using linear SVR^25^, suggesting that a linear model averaging over many patients may approximate the same surface a nonlinear model captures directly in smaller samples. The small aggregate imaging gain (ΔF1 ≤ 0.029) may be read as indicating that imaging is expendable, but the specific anatomical pattern observed (shared core, ventral-dominant resolution, dorsal-plus-interhemispheric discourse) is the one predicted by the literature prior to model fitting, suggesting that mechanistic interpretation and aggregate classification reward different kinds of evidence. Finally, both final models systematically overpredicted 12-month scores for patients who remained severely aphasic. In the n = 8 patients with WAB-AQ_12mo_ < 50, the FS4 RF model overpredicted WAB-AQ by a mean of +10.5 points (MAE = 17.2); the analogous pattern was present for discourse output (n = 11 with CU_12mo_ < 10; mean overprediction = +8.8 content units, MAE = 9.0), a well-documented regression-to-the-mean artifact in aphasia outcome prediction^8,52^. Adaptive designs that re-stratify patients at intermediate timepoints (e.g., 3 months) based on deviation from predicted trajectories may be required to reach this subgroup, and multi-site collaboration will be needed to accumulate sufficient severe cases for low-severity-specific training.

### Limitations

Several limitations warrant consideration. First, the single-site n = 73 limits generalizability, though the sample is comparable to other ML aphasia prediction studies. Second, CUs index only one dimension of discourse; fluency, grammatical complexity, and communicative efficiency are not captured^26,28^, and the more modest discourse model performance indicates that substantial discourse-relevant variance lies outside the features and the outcome sampled here. Third, SLP receipt tracked acute severity (52 of 73 patients [71.2%] received SLP, presenting with more severe acute aphasia than non-recipients: 62.2 ± 36.4 vs 93.7 ± 12.4 WAB-AQ, p < .001; Supplement S8), consistent with the clinical reality that patients with more severe acute aphasia typically receive more intensive therapy. A supplementary sensitivity analysis (Supplement S8) found no systematic association between treatment exposure and residual prediction error, suggesting that the predictor rankings reported here are not confounded by differential therapy allocation; however, the observational design cannot fully rule out treatment effects on recovery trajectories, consistent with Wilson et al.^2^. Fourth, the observational design captures in part spontaneous recovery and cannot disentangle prediction from treatment effects, so integrating prediction with randomized allocation remains a future step. Pairing acute-phase prediction with emerging digital twin frameworks, which simulate individual treatment responses and the effect of modifiable health factors on recovery, offers a synergistic path toward personalizing rehabilitation^53,54^.

### Conclusions

Machine learning models trained on acute-phase data predict 12-month aphasia resolution with high accuracy and discourse content normalization with more modest accuracy. Clinical features carry most of the predictive variance for aphasia resolution, whereas imaging contributes modestly more for discourse normalization and, for both outcomes, provides an interpretive window onto the shared and divergent neural substrates of recovery. Clinical translation will require prospective external validation and integration into point-of-care-like tools, e.g., a bedside risk calculator that uses acute WAB-AQ, lesion volume, and key imaging features and returns a 12-month resolution probability, enabling early intensive intervention for patients at highest risk of persistent aphasia.

## Data availability

The code used for all analyses is publicly available in an Open Science Framework repository (https://osf.io/m8uv2/overview?view_only=0e0f6c5e75cd4f198c50fa23c3e34a16). Our data are available from the Vivli controlled repository using the Sponsor Protocol ID: NA_00042097 https://doi.org/10.25934/PR00012002. The identifiable data may not be shared publicly, as they represent clinical protected health information in participants who did not consent to have their data banked. Currently, data are available upon request to the authors, subject to review by the Johns Hopkins University School of Medicine Institutional Review Board (contact via) resulting in a formal data sharing agreement.

## Supporting information

Checklist

## Acknowledgements

The authors sincerely thank the study participants and their families for their generous contribution of time and data, as well as research coordinators, speech-language pathologists who contributed to acute assessments, and our imaging technologists.

