## Supplementary material for "Acute-Phase Machine Learning Prediction of 12-Month Aphasia and Discourse Recovery": Checklist

### TRIPOD+AI Checklist

Reference: Collins GS, Moons KGM, Dhiman P, et al. TRIPOD+AI statement: updated guidance for reporting clinical prediction models that use regression or machine learning methods. BMJ. 2024;385:e078378. doi:10.1136/bmj-2023-078378

| Section / Topic | Item | D/E | Checklist item | Reported in |
|---|---|---|---|---|
| **TITLE** | | | | |
| **Title** | 1 | D;E | Identify the study as developing or evaluating the performance of a multivariable prediction model, the target population, and the outcome to be predicted | Title page |
| **ABSTRACT** | | | | |
| **Abstract** | 2 | D;E | See TRIPOD+AI for Abstracts checklist | Abstract |
| **INTRODUCTION** | | | | |
| **Background** | 3a | D;E | Explain the healthcare context (including whether diagnostic or prognostic) and rationale for developing or evaluating the prediction model, including references to existing models | Introduction, ¶1–5 |
| **Background** | 3b | D;E | Describe the target population and the intended purpose of the prediction model in the context of the care pathway, including its intended users (e.g., healthcare professionals, patients, public) | Introduction, ¶1 and ¶7 (clinical translation framing); Discussion, Conclusions (bedside risk-calculator use case) |
| **Background** | 3c | D;E | Describe any known health inequalities between sociodemographic groups | Not directly add<br>ressed; noted as a limitation in Discussion > Limitations (single-site cohort; generalizability constraint) |
| **Objectives** | 4 | D;E | Specify the study objectives, including whether the study describes the development or validation of a prediction model (or both) | Introduction, ¶7 (aims); Development only, no external validation in this study |
| **METHODS** | | | | |
| **Data** | 5a | D;E | Describe the sources of data separately for the development and evaluation datasets | Materials and methods > Participants (Johns Hopkins Hospital acute stroke cohort, 2007–2024) |

| | | | | |
|---|---|---|---|---|
| | | | (e.g., randomised trial, cohort, routine care or registry data), the rationale for using these data, and representativeness of the data | |
| **Data** | 5b | D;E | Specify the dates of the collected participant data, including start and end of participant accrual; and, if applicable, end of follow-up | Materials and methods > Participants (accrual 2007–2024; 12-month follow-up) |
| **Participants** | 6a | D;E | Specify key elements of the study setting (e.g., primary care, secondary care, general population) including the number and location of centres | Materials and methods > Participants (single-centre tertiary stroke center, Baltimore, MD, USA) |
| **Participants** | 6b | D;E | Describe the eligibility criteria for study participants | Materials and methods > Participants (inclusion criteria explicit) |
| **Participants** | 6c | D;E | Give details of any treatments received, and how they were handled during model development or evaluation, if relevant | Discussion > Limitations (SLP receipt tracked acute severity); Supplementary S8 (treatment-effects sensitivity analysis) |
| **Data preparation** | 7 | D;E | Describe any data pre-processing and quality checking, including whether this was similar across relevant sociodemographic groups | Materials and methods > Neuroimaging Processing (OpenADS segmentation, blinded manual review); Materials and methods > Machine Learning Pipeline (within-fold standardization, volume residualization, inverse-density weighting) |
| **Outcome** | 8a | D;E | Clearly define the outcome that is being predicted and the time horizon, including how and when assessed, the rationale for choosing this outcome, and whether the method of outcome assessment is consistent across sociodemographic groups | Materials and methods > Outcome Measures (WAB-AQ ≥ 93.8 for aphasia resolution; Modern Cookie Theft content units ≥ 22.1 for discourse normalization; both at 12 months) |
| **Outcome** | 8b | D;E | If outcome assessment requires subjective interpretation, describe the qualifications and demographic characteristics of the outcome assessors | Materials and methods > Outcome Measures (standardized WAB-R administration); content-unit scoring described in Berube et al., 2019 performed on the same population |
| **Outcome** | 8c | D;E | Report any actions to blind assessment of the outcome to be predicted | Materials and methods > Neuroimaging Processing (lesion masks reviewed by research assistant blinded to 12-month outcome) |
| **Predictors** | 9a | D | Describe the choice of initial predictors (e.g., literature, previous models, all available | Materials and methods > Feature Sets (hierarchical FS1–FS4 defined a priori from literature and dual-stream model); Supplementary S1–S3 (selection rationales) |

| | | | | |
|---|---|---|---|---|
| | | | predictors) and any pre-selection of predictors before model building | |
| **Predictors** | 9b | D;E | Clearly define all predictors, including how and when they were measured (and any actions to blind assessment of predictors for the outcome and other predictors) | Materials and methods > Feature Sets; Supplementary Table S3 (complete feature list); acute WAB-AQ assessed within first week post-onset |
| **Predictors** | 9c | D;E | If predictor measurement requires subjective interpretation, describe the qualifications and demographic characteristics of the predictor assessors | Materials and methods > Neuroimaging Processing (manual lesion mask correction by trained research assistant, verified by lead author, both blinded to outcome) |
| **Sample size** | 10 | D;E | Explain how the study size was arrived at (separately for development and evaluation), and justify that the study size was sufficient to answer the research question. Include details of any sample size calculation | Materials and methods > Participants (convenience cohort); Discussion > Limitations (n = 73 discussed relative to comparable ML aphasia prediction studies) |
| **Missing data** | 11 | D;E | Describe how missing data were handled. Provide reasons for omitting any data | Materials and methods > Outcome Measures (discourse outcome N = 61 reflects subset with completed 12-month Modern Cookie Theft sampling); complete-case approach; Table 1 comparisons |
| **Analytical methods** | 12a | D | Describe how the data were used (e.g., for development and evaluation of model performance) in the analysis, including whether the data were partitioned, considering any sample size requirements | Materials and methods > Machine Learning Pipeline (nested 10-fold × 10-repeat cross-validation; outer folds stratified on outcome quartiles) |
| **Analytical methods** | 12b | D | Depending on the type of model, describe how predictors were handled in the analyses (functional form, rescaling, transformation, or any standardisation) | Materials and methods > Machine Learning Pipeline (within-fold z-score standardization; imaging features residualized against lesion volume) |
| **Analytical methods** | 12c | D | Specify the type of model, rationale, all model-building steps, including any hyperparameter tuning, and method for internal validation | Materials and methods > Machine Learning Pipeline (Ridge, SVR, RF, XGBoost); Supplementary S4 (algorithm rationale); Supplementary Table S1 (hyperparameter grids) |
| **Analytical methods** | 12d | D;E | Describe if and how any heterogeneity in estimates of model parameter values and | Not applicable (single-site cohort) |

| | | | | |
|---|---|---|---|---|
| | | | model performance was handled and quantified across clusters (e.g., hospitals, countries) | |
| **Analytical methods** | 12e | D;E | Specify all measures and plots used (and their rationale) to evaluate model performance (e.g., discrimination, calibration, clinical utility) and, if relevant, to compare multiple models | Materials and methods > Model Evaluation (Pearson r, $R^2$, MAE, F1, sensitivity, specificity, PPV, NPV, balanced accuracy, MCC); Supplementary S6 (metric definitions and rationale); Figures 2–3 (prediction plots) |
| **Analytical methods** | 12f | E | Describe any model updating (e.g., recalibration) arising from the model evaluation, either overall or for particular sociodemographic groups or settings | Not applicable (development study; no updating performed) |
| **Analytical methods** | 12g | E | For model evaluation, describe how the model predictions were calculated (e.g., formula, code, object, application programming interface) | Code available at OSF repository (https://osf.io/m8uv2/?view_only=0e0f6c5e75cd4f198c50fa23c3e34a16; see Data Availability) |
| **Class imbalance** | 13 | D;E | If class imbalance methods were used, state why and how this was done, and any subsequent methods to recalibrate the model or the model predictions | Materials and methods > Machine Learning Pipeline (inverse-density sample weighting for left-skewed outcomes); Supplementary S5 (KDE parameters and rationale) |
| **Fairness** | 14 | D;E | Describe any approaches that were used to address model fairness and their rationale | Not formally evaluated in the present study; Discussion > Limitations acknowledges single-site cohort and generalizability constraints |
| **Model output** | 15 | D | Specify the output of the prediction model (e.g., probabilities, classification). Provide details and rationale for any classification and how the thresholds were identified | Materials and methods > Outcome Measures and Machine Learning Pipeline (regression-then-threshold design; clinical cutoffs WAB-AQ ≥ 93.8 and CU ≥ 22.1 applied post-hoc to continuous predictions) |
| **Training vs evaluation** | 16 | D;E | Identify any differences between the development and evaluation data in healthcare setting, eligibility criteria, outcome, and predictors | Not applicable (internal cross-validation only; no separate evaluation dataset) |
| **Ethical approval** | 17 | D;E | Name the institutional research board or ethics committee that approved the study and describe the | Johns Hopkins Medicine IRB protocol NA_00042097; written informed consent obtained from all participants in accordance with the Declaration of Helsinki, see Materials and methods > Participants |

| | | | | |
|---|---|---|---|---|
| | | | participant-informed consent or the ethics committee waiver of informed consent | |
| **OPEN SCIENCE** | | | | |
| **Funding** | 18a | D;E | Give the source of funding and the role of the funders for the present study | Funding section |
| **Conflicts of interest** | 18b | D;E | Declare any conflicts of interest and financial disclosures for all authors | Competing interests section |
| **Protocol** | 18c | D;E | Indicate where the study protocol can be accessed or state that a protocol was not prepared | No formal pre-registered protocol; analysis plan and feature sets were specified a priori (Materials and methods > Feature Sets) |
| **Registration** | 18d | D;E | Provide registration information for the study, including register name and registration number, or state that the study was not registered | Not registered (observational development study) |
| **Data sharing** | 18e | D;E | Provide details of the availability of the study data | Data availability section |
| **Code sharing** | 18f | D;E | Provide details of the availability of the analytical code | Data Availability section (OSF: https://osf.io/m8uv2/?view_only=0e0f6c5e75cd4f198c50fa23c3e34a16) |
| **PATIENT & PUBLIC INVOLVEMENT** | | | | |
| **Patient & Public Involvement** | 19 | D;E | Provide details of any patient and public involvement during the design, conduct, reporting, interpretation, or dissemination of the study or state no involvement | No formal patient or public involvement in study design or analysis |
| **RESULTS** | | | | |
| **Participants** | 20a | D;E | Describe the flow of participants through the study, including the number of participants with and without the outcome and, if applicable, a summary of the follow-up time. A diagram may be helpful | Materials and methods > Participants; Results > Aphasia Resolution (n = 73: 47 resolved, 26 not); Results > Discourse Content Normalization (n = 61: 28 normalized, 33 not) |
| **Participants** | 20b | D;E | Report the characteristics overall and, where applicable, for each data source or setting, including the key dates, key predictors (including demographics), treatments received, sample size, number of outcome events, follow-up time, and amount of | Table 1 (participant characteristics with between-group comparisons) |

| | | | | |
|---|---|---|---|---|
| | | | missing data. A table may be helpful. Report any differences across key demographic groups | |
| **Participants** | 20c | E | For model evaluation, show a comparison with the development data of the distribution of important predictors (demographics, predictors, and outcome) | Not applicable (no separate evaluation dataset) |
| **Model development** | 21 | D;E | Specify the number of participants and outcome events in each analysis (e.g., for model development, hyperparameter tuning, model evaluation) | Results; Table 2 (performance per outcome); Materials and methods > Machine Learning Pipeline (nested 10×10 CV) |
| **Model specification** | 22 | D | Provide details of the full prediction model (e.g., formula, code, object, application programming interface) to allow predictions in new individuals and to enable third-party evaluation and implementation, including any restrictions to access or re-use | Code and trained-model objects available at OSF repository (https://osf.io/m8uv2/?view_only=0e0f6c5e75cd4f198c50fa23c3e34a16; see Data Availability) |
| **Model performance** | 23a | D;E | Report model performance estimates with confidence intervals, including for any key subgroups (e.g., sociodemographic). Consider plots to aid presentation | Table 2 (all metrics with 95% bootstrap CIs); Figures 2–3; Supplementary Table S2 (complete leaderboard for all 32 combinations) |
| **Model performance** | 23b | D;E | If examined, report results of any heterogeneity in model performance across clusters | Not applicable (single site) |
| **Model updating** | 24 | E | Report the results from any model updating, including the updated model and subsequent performance | Not applicable (no updating performed) |
| **DISCUSSION** | | | | |
| **Interpretation** | 25 | D;E | Give an overall interpretation of the main results, including issues of fairness in the context of the objectives and previous studies | Discussion ¶1–6 |
| **Limitations** | 26 | D;E | Discuss any limitations of the study (such as a non-representative sample, sample size, overfitting, missing | Discussion > Limitations |

| | | | | |
|---|---|---|---|---|
| | | | data) and their effects on any biases, statistical uncertainty, and generalizability | |
| **Usability** | 27a | D | Describe how poor quality or unavailable input data (e.g., predictor values) should be assessed and handled when implementing the prediction model | Conclusions (bedside risk-calculator) |
| **Usability** | 27b | D | Specify whether users will be required to interact in the handling of the input data or use of the model, and what level of expertise is required of users | Conclusions (intended for clinician-facing bedside use; WAB-AQ administration requires standard SLP/neurology expertise) |
| **Usability** | 27c | D;E | Discuss any next steps for future research, with a specific view to applicability and generalizability of the model | Discussion > Limitations (prospective external validation; adaptive designs for severe subgroup; digital-twin integration) |
