## Supplemental Materials for "Acute-Phase Machine Learning Prediction of 12-Month Aphasia and Discourse Recovery"

### Supplementary Materials

#### Abbreviations

AG = angular gyrus; BA = Brodmann area; BPM = Brain Parcellation Map; CU = content units; CV = cross-validation; FS1–FS4 = feature sets 1 through 4; FUS = fusiform gyrus; IFG = inferior frontal gyrus; IFOF = inferior fronto-occipital fasciculus; ILF = inferior longitudinal fasciculus; INS = insula; IPG = inferior parietal gyrus; ITG = inferior temporal gyrus; KDE = kernel density estimation; L/R = left/right; LH = left hemisphere; MAE = mean absolute error; MCC = Matthews Correlation Coefficient; MFG = middle frontal gyrus; MTG = middle temporal gyrus; NeMo = Network Modification; NPV = negative predictive value; POP = pars opercularis; PPV = positive predictive value; PrCG = precentral gyrus; PTR = pars triangularis; PU = putamen; RBF = radial basis function; RF = random forest; RFE = recursive feature elimination; SFG = superior frontal gyrus; SHAP = SHapley Additive exPlanations; SLF = superior longitudinal fasciculus; SLP = speech-language pathology; SMG = supramarginal gyrus; STG = superior temporal gyrus; SVR = support vector regression; TH = thalamus; UF = uncinate fasciculus; WAB = Western Aphasia Battery–Revised Aphasia Quotient; XGBoost = extreme gradient boosting.

#### Supplemental Methods

*S1. Cortical Region Selection Rationale*

The 10 LH cortical regions in FS3/FS4 were selected a priori from the BPM atlas[1] based on convergent evidence from lesion-symptom mapping[2] and the dual-stream model[3]. Eight regions fall within canonical left perisylvian language territory with documented lesion-behavior associations to production, comprehension, or repetition: IFG pars opercularis (BA 44) and pars triangularis (BA 45) for speech production and syntactic processing[4] ; STG for auditory comprehension via the ventral phonological-recognition pathway[2]; SMG and angular gyrus for phonological and lexical-semantic processing[2]; MTG and ITG for lexical-semantic retrieval[2]; and insula for speech motor planning[2,5]. Two extra-perisylvian frontal regions were included to test contributions outside the core perisylvian network: MFG, given its role in executive-control aspects of connected-speech production[2], and SFG, as the cortical origin of the frontal aslant tract supporting speech initiation and verbal fluency[6], with associations to complex syntactic processing[7]. Subcortical structures (thalamus, putamen) were excluded from the cortical-parcel feature class to avoid conflating distinct tissue types; thalamic contributions are instead indexed by the NeMo L.thalamus–POP connectivity feature (see S3).

*S2. White Matter Tract Selection*

Five LH tracts from Tractotron[8] were selected to span the dual-stream model[3,9]: dorsal stream (SLF, arcuate fasciculus) for phonological-articulatory processing, and ventral stream (IFOF, ILF, UF) for semantic access and comprehension. Tract selection was restricted to pathways with an established role in language per the dual-stream architecture; commissural, motor, and non-language association tracts were not included. Right-hemisphere tract homologues were not modeled as tract features because all lesions were left-hemisphere; interhemispheric structural influence is instead captured at the network level via the NeMo connectivity features (see S3). Disconnection probability (0–1) quantifies the likelihood that each tract is disrupted by the lesion[8].

*S3. NeMo Language Circuit Selection*

Twenty-two pairwise NeMo Change-in-Connectivity (ChaCo) scores[10,11] were defined a priori to index canonical language circuits within the fs86 Desikan-Killiany parcellation[12]: dorsal stream core (POP–STG, POP–SMG, PTR–STG, POP–MTG, PTR–MTG), receptive network (SMG–STG, IPG–STG, ITG–STG, AG–STG), production network (POP–PTR, INS–POP, INS–STG, SFG–POP, L.thalamus–POP), interhemispheric homologues (L.SMG–R.SMG, L.STG–R.STG, L.POP–R.POP, L.PTR–R.PTR, L.SFG–R.SFG), and extended semantic circuits (AG–MTG, MTG–ITG, FUS–ITG). Pairs were specified before model fitting. The interhemispheric homologue set indexes the hypothesis that preserved cross-hemispheric connectivity between homologous language nodes supports later right-hemisphere recruitment during recovery, a process that is regionally heterogeneous across homotopic nodes[13]. This hypothesis is separable from within-LH tract integrity (S2): the former tests a structural precondition for cross-hemispheric compensation, while the latter tests preserved within-network pathway integrity. All 43 features in FS4 (5 FS1 clinical + 1 FS2 lesion volume + 10 FS3 cortical parcels + 5 FS3 white matter tracts + 22 FS4 NeMo pairs) are listed in Supplementary Table S3.

*S3a. Model Complexity*

The FS4 specification includes 43 features in a cohort of 73 patients (61 for discourse), yielding a feature-to-sample ratio below the 10:1 heuristic sometimes invoked for classical regression[14]. Four design choices mitigate the associated overfitting risk. First, the hierarchical feature-set scheme (FS1–FS4) allows direct comparison of parsimonious versus expanded specifications on the same cohort, i.e., any gain from imaging features must exceed the penalty of added parameters (observed $\Delta F1 \leq 0.029$ from FS1 to FS4; Supplementary Table S2). Second, all preprocessing, feature residualization, and hyperparameter tuning were performed inside training folds only, precluding test-fold information from influencing model selection[15,16]. Third, regularized (Ridge, SVR) and tree-ensemble (RF, XGBoost) learners each incorporate intrinsic model-complexity control via L2 penalties, kernel regularization, tree-depth constraints, and early stopping. Fourth, nested cross-validation with a 10-fold outer × 10-repeat resampling scheme provides out-of-fold generalization estimates that are insulated from single-partition idiosyncrasies (Supplementary Table S1).

*S4. Algorithm Justification*

The four algorithms were selected to span the linear–nonlinear and single-model–ensemble axes of the ML landscape. Ridge regression[17] provides a regularized linear baseline that stabilizes coefficient estimates under correlated imaging features. Support vector regression with a radial basis function kernel[18] captures nonlinear structure in the severity–outcome surface and was the top-performing model in Hu et al.[19] for chronic aphasia severity prediction from multimodal imaging. Random forest[20] accommodates high-order feature interactions without explicit specification and is robust to outliers and to mixed feature scales. XGBoost[21] extends tree-based learning with gradient boosting and sequential error correction, providing an ensemble counterpart to RF that is tuned for smaller datasets with structured feature dependencies.

*S5. Inverse-Density Sample Weighting*

Twelve-month outcome distributions for both WAB-AQ (N = 73) and content units (N = 61) were left-skewed. Most patients reached or approached ceiling, while the tail of poor-outcome cases was sparse. Under standard loss functions this asymmetry allows models to optimize performance on the majority at the expense of patients with poor outcomes, precisely the subgroup of greatest clinical interest. Density-based reweighting counteracts this bias by increasing the effective loss contribution of under-represented cases[22].

***KDE parameters.*** Bandwidth was set by Silverman's rule of thumb[23] and capped at a maximum of 5 points on the 0–100 outcome scale to prevent over-smoothing in the low-density tail. For each subject i with outcome $y_i$, the pre-normalization weight was set proportional to 1 / KDE($y_i$). Weights were mean-normalized to 1.0 across the training fold to preserve effective sample size, then clipped at 5× the median weight to prevent any individual low-outcome case from dominating the loss.

*S6. Evaluation Metrics: Definitions and Rationale*

**Continuous-fit metrics.** Pearson r quantifies the linear association between predicted and observed 12-month scores. $R^2$ is the coefficient of determination, i.e., the proportion of observed-score variance accounted for by the model; $R^2 < 0$ indicates worse-than-mean performance. MAE is the mean absolute error, reported in WAB-AQ points (aphasia resolution) or content units (discourse). These three metrics collectively evaluate both the rank-order and the absolute calibration of continuous predictions[24].

**Binary-classification metrics.** Predictions were dichotomized by applying the clinical threshold (WAB-AQ ≥ 93.8 or CU ≥ 22.1) to each subject's predicted continuous score. From the resulting confusion matrix (TP, TN, FP, FN), the following derived metrics were computed: sensitivity (TP / [TP + FN]), specificity (TN / [TN + FP]), positive predictive value (PPV = TP / [TP + FP]), negative predictive value (NPV = TN / [TN + FN]), F1 (harmonic mean of PPV and sensitivity[25]), balanced accuracy (mean of sensitivity and specificity[26]), and Matthews Correlation Coefficient (MCC[27]), a single-number summary of the full confusion matrix ranging from −1 to +1, with 0 corresponding to chance at any base rate.

**Base-rate benchmarks.** Under the 64.4% aphasia-resolution base rate, a naive all-positive classifier would achieve raw accuracy = 0.644, balanced accuracy = 0.500, and MCC = 0.000. F1 alone can be misleading under class imbalance, hence we report MCC and balanced accuracy alongside F1 to adjust for this.

**Rationale for F1 as the primary selection metric.** F1 was chosen because the clinical question is identification of patients unlikely to resolve, a problem where both false negatives (missed at-risk patients) and false positives (over-triage of low-risk patients to high-intensity intervention) carry meaningful cost. F1 penalizes asymmetry in either direction. F1 was also the primary metric in comparable recent ML aphasia recovery studies[19,28], enabling direct cross-study comparison.

*Uncertainty quantification.* 95% bootstrap confidence intervals were computed from 2,000 resamples of the out-of-fold predictions using the percentile method[29]. Permutation-based P values were computed by shuffling outcome labels prior to model fitting and re-running the full nested cross-validation pipeline 1,000 times[30]; the permutation P value is the proportion of shuffled runs achieving F1 ≥ the observed model's F1. Reporting follows the TRIPOD+AI guideline[31].

*S7. SHAP Analysis: Computation and Stability Protocol*

SHAP values (SHapley Additive exPlanations**[32]**) decompose a single prediction into per-feature contributions, quantifying how much each feature shifted the model's output for that subject relative to the mean prediction. The Shapley-value formulation is drawn from cooperative game theory and averages each feature's marginal contribution over all possible feature coalitions. For the FS4 random forest model (aphasia resolution), TreeSHAP was used to compute exact Shapley decompositions on each test-fold tree ensemble. For the FS4 SVR model (discourse content), KernelSHAP was used with background samples drawn from the training fold.

**Stability protocol**. Per-subject SHAP values were computed for every test-fold prediction across all 100 outer folds (10 folds × 10 repeats). For each feature, per-fold importance was summarized as the mean absolute SHAP value (mean |SHAP|) across that fold's test subjects. Feature rankings reported in the main text (Table 3) and in Supplementary Figure S1 were then obtained by averaging fold-level mean |SHAP| across all 100 outer folds. This across-fold averaging provides stability against idiosyncratic within-fold noise.

**Continuous-score decomposition**. Because the pipeline was constructed as regression-then-threshold[14,33], SHAP decomposes each subject's continuous-score prediction. The same decomposition therefore informs both the binary classification result (whether the subject crossed the clinical threshold) and the continuous fit (how close the prediction was to the observed 12-month score), avoiding the need for a separate classification-side SHAP computation with its associated recalibration assumptions.

*S8. Treatment-Effects Sensitivity Check*

To evaluate whether the predictor rankings reported in the main text were influenced by post-stroke therapy exposure, we conducted a sensitivity analysis using a binary indicator of whether the patient received any formal speech-language pathology (SLP) services between acute hospitalization and 12-month follow-up (yes/no), recorded from medical records and patient self-report.

Cohort composition. Of the 73 patients, 52 (71.2%) received SLP and 21 (28.8%) did not. Consistent with typical clinical referral patterns, patients who received SLP had more severe acute aphasia (62.17 ± 36.39 vs 93.74 ± 12.42 WAB-AQ, $P < 0.001$) and larger lesions

(49.54 ± 74.45 vs 4.79 ± 4.80 mL, P < 0.001). The two groups did not differ on demographic variables (age, sex, education, prior-stroke history; Mann-Whitney U and Fisher exact tests; all P > 0.76).

**Analysis**. For each final model (FS4 RF for aphasia resolution; FS4 SVR for discourse normalization), prediction residuals (observed − predicted 12-month score) were tested for association with SLP receipt using Mann-Whitney U tests (Mann & Whitney, 1947). In a secondary analysis, we re-aggregated each model's per-subject absolute SHAP values over the SLP-recipient subgroup and compared the resulting top-15 feature rankings to those from the full cohort using Spearman rank correlation. This approach avoids re-training models on the smaller and baseline-imbalanced subgroup while still isolating feature importance as learned about treated patients specifically.

**Results**. Residuals did not differ significantly between SLP recipients and non-recipients for either outcome (aphasia resolution: median residual −1.0 vs −0.3 WAB-AQ points, P = 0.221; discourse normalization: median residual 3.3 vs 5.6 content units, P = 0.541). The top-15 SHAP rankings computed over the SLP-recipient subgroup correlated strongly with those from the full cohort for both outcomes (aphasia resolution: Spearman ρ = 0.99, P < 0.001; discourse normalization: Spearman ρ = 0.98, P < 0.001), with shared core features (acute WAB-AQ, lesion volume, left pars triangularis) retaining their top-10 positions in both analyses.

This pattern is consistent with Wilson et al.[34], who reported that treatment exposure did not meaningfully alter the ranking of recovery predictors in an acute-to-chronic aphasia cohort. These results are necessarily limited by the observational nature of treatment allocation. The SLP-receipt binary does not capture dose, frequency, or modality, and patients with more severe initial aphasia are typically more likely to receive therapy (as reflected in the baseline imbalance reported above). These results should not be interpreted as evidence that treatment does not influence recovery, only that treatment exposure as operationalized here is not confounded with the SHAP-identified predictor rankings.

### Supplemental Tables

*Supplementary Table S1. Hyperparameter Grids and Cross-Validation Settings.*

| Model | Hyperparameters (grid searched) |
|---|---|
| Ridge | alpha: {0.001, 0.01, 0.1, 1, 10, 100, 1000} |
| SVR | C: {0.1, 1, 10, 100}; gamma: {0.001, 0.01, 0.1, 1}; epsilon: {0.1, 0.5, 1} |
| RF | max_depth: {None, 5, 10}; min_samples_leaf: {1, 2, 4}; max_features: {0.5, sqrt, log2}; n_estimators = 200 |
| XGBoost | n_estimators: {50, 100, 200}; max_depth: {3, 5, 7}; learning_rate: {0.01, 0.05, 0.1}; subsample: {0.6, 0.8} |


| Model | Hyperparameters (grid searched) |
|---|---|
| Ridge | alpha: {0.001, 0.01, 0.1, 1, 10, 100, 1000} |
| SVR | C: {0.1, 1, 10, 100}; gamma: {0.001, 0.01, 0.1, 1}; epsilon: {0.1, 0.5, 1} |
| RF | max_depth: {None, 5, 10}; min_samples_leaf: {1, 2, 4}; max_features: {0.5, sqrt, log2}; n_estimators = 200 |
| XGBoost | n_estimators: {50, 100, 200}; max_depth: {3, 5, 7}; learning_rate: {0.01, 0.05, 0.1}; subsample: {0.6, 0.8} |


*Note:* Nested CV: 10-fold × 10 repeats (outer), 10-fold GridSearchCV (inner). RFE (step = 0.1) applied to FS3/FS4.

*Supplementary Table S2. Complete Model Performance Leaderboard (all 32 combinations) with 95% Bootstrap Confidence Intervals.*

| **Outcome** | **FS** | **Model** | **F1 [95% CI]** | **BalAcc [95% CI]** | **MCC [95% CI]** | **Sens [95% CI]** | **Spec [95% CI]** | **Pearson r [95% CI]** | **$R^2$ [95% CI]** | **MAE [95% CI]** |
|---|---|---|---|---|---|---|---|---|---|---|
| WAB | FS2 | RF | 0.874 [0.800–0.941] | 0.866 [0.787–0.938] | 0.704 [0.548–0.842] | 0.809 [0.700–0.913] | 0.923 [0.818–1.000] | 0.824 [0.674–0.936] | 0.647 [0.149–0.871] | 7.32 [4.60–10.21] |

| Outcome | FS | Model | F1 [95% CI] | BalAcc [95% CI] | MCC [95% CI] | Sens [95% CI] | Spec [95% CI] | Pearson r [95% CI] | R² [95% CI] | MAE [95% CI] |
|---|---|---|---|---|---|---|---|---|---|---|
| WAB | FS3 | RF | 0.874 [0.800–0.941] | 0.866 [0.787–0.938] | 0.704 [0.548–0.842] | 0.809 [0.700–0.913] | 0.923 [0.818–1.000] | 0.831 [0.687–0.937] | 0.661 [0.178–0.870] | 7.28 [4.62–10.02] |
| **WAB** | **FS4** | **RF** | **0.874 [0.800–0.941]** | **0.866 [0.787–0.938]** | **0.704 [0.548–0.842]** | **0.809 [0.700–0.913]** | **0.923 [0.818–1.000]** | **0.827 [0.678–0.937]** | **0.652 [0.154–0.873]** | **7.26 [4.56–10.06]** |
| WAB | FS2 | Ridge | 0.871 [0.790–0.943] | 0.874 [0.802–0.938] | 0.718 [0.572–0.844] | 0.787 [0.667–0.896] | 0.962 [0.880–1.000] | 0.778 [0.662–0.880] | 0.532 [-0.021–0.760] | 9.65 [6.57–12.56] |
| WAB | FS2 | SVR | 0.867 [0.780–0.936] | 0.838 [0.742–0.923] | 0.658 [0.470–0.832] | 0.830 [0.721–0.930] | 0.846 [0.690–1.000] | 0.800 [0.655–0.906] | 0.582 [0.004–0.809] | 8.66 [5.74–11.58] |
| WAB | FS4 | SVR | 0.854 [0.763–0.930] | 0.827 [0.731–0.912] | 0.634 [0.444–0.796] | 0.809 [0.700–0.913] | 0.846 [0.690–1.000] | 0.795 [0.652–0.904] | 0.586 [0.040–0.801] | 8.64 [5.66–11.48] |
| WAB | FS1 | RF | 0.851 [0.776–0.921] | 0.836 [0.743–0.915] | 0.646 [0.470–0.802] | 0.787 [0.682–0.894] | 0.885 [0.750–1.000] | 0.832 [0.686–0.941] | 0.662 [0.167–0.876] | 7.18 [4.57–10.00] |
| WAB | FS2 | XGBoost | 0.847 [0.771–0.921] | 0.845 [0.770–0.920] | 0.660 [0.501–0.809] | 0.766 [0.660–0.875] | 0.923 [0.818–1.000] | 0.800 [0.652–0.926] | 0.585 [-0.005–0.842] | 8.43 [5.65–11.45] |

| Outcome | FS | Model | F1 [95% CI] | BalAcc [95% CI] | MCC [95% CI] | Sens [95% CI] | Spec [95% CI] | Pearson r [95% CI] | R² [95% CI] | MAE [95% CI] |
|---|---|---|---|---|---|---|---|---|---|---|
| WAB | FS3 | Ridge | 0.843 [0.761–0.920] | 0.853 [0.780–0.924] | 0.676 [0.521–0.821] | 0.745 [0.630–0.857] | 0.962 [0.880–1.000] | 0.776 [0.664–0.877] | 0.530 [-0.031–0.754] | 9.73 [6.57–12.64] |
| WAB | FS3 | SVR | 0.841 [0.747–0.917] | 0.817 [0.719–0.908] | 0.611 [0.418–0.771] | 0.787 [0.667–0.894] | 0.846 [0.690–1.000] | 0.808 [0.670–0.903] | 0.613 [0.135–0.805] | 8.65 [5.85–11.33] |
| WAB | FS3 | XGBoost | 0.833 [0.750–0.911] | 0.834 [0.754–0.911] | 0.640 [0.480–0.792] | 0.745 [0.634–0.857] | 0.923 [0.818–1.000] | 0.815 [0.683–0.928] | 0.616 [0.126–0.840] | 8.36 [5.65–11.19] |
| WAB | FS4 | Ridge | 0.829 [0.741–0.905] | 0.842 [0.772–0.910] | 0.657 [0.505–0.791] | 0.723 [0.595–0.840] | 0.962 [0.880–1.000] | 0.778 [0.653–0.885] | 0.540 [-0.018–0.764] | 9.47 [6.39–12.40] |
| WAB | FS4 | XGBoost | 0.805 [0.718–0.889] | 0.813 [0.732–0.896] | 0.599 [0.438–0.758] | 0.702 [0.580–0.830] | 0.923 [0.818–1.000] | 0.810 [0.671–0.926] | 0.609 [0.099–0.838] | 8.37 [5.69–11.23] |
| WAB | FS1 | Ridge | 0.737 [0.611–0.842] | 0.779 [0.693–0.860] | 0.545 [0.370–0.681] | 0.596 [0.449–0.739] | 0.962 [0.870–1.000] | 0.763 [0.643–0.849] | 0.471 [-0.157–0.705] | 12.68 [9.94–15.38] |
| WAB | FS1 | XGBoost | 0.737 [0.620–0.840] | 0.779 [0.694–0.856] | 0.545 [0.391–0.697] | 0.596 [0.465–0.729] | 0.962 [0.880–1.000] | 0.759 [0.604–0.890] | 0.534 [-0.026–0.765] | 9.71 [6.83–12.71] |

| Outcome | FS | Model | F1 [95% CI] | BalAcc [95% CI] | MCC [95% CI] | Sens [95% CI] | Spec [95% CI] | Pearson r [95% CI] | R² [95% CI] | MAE [95% CI] |
|---|---|---|---|---|---|---|---|---|---|---|
| WAB | FS1 | SVR | 0.685 [0.537–0.795] | 0.747 [0.662–0.818] | 0.493 [0.333–0.622] | 0.532 [0.381–0.673] | 0.962 [0.870–1.000] | 0.743 [0.619–0.838] | 0.440 [-0.185–0.677] | 12.32 [9.38–15.18] |
| **NCT** | **FS4** | **SVR** | **0.725 [0.593–0.831]** | **0.704 [0.602–0.800]** | **0.433 [0.220–0.647]** | **0.893 [0.767–1.000]** | **0.515 [0.343–0.684]** | **0.617 [0.437–0.753]** | **0.252 [-0.164–0.512]** | **8.96 [7.39–10.64]** |
| NCT | FS1 | Ridge | 0.704 [0.571–0.806] | 0.674 [0.577–0.773] | 0.380 [0.164–0.570] | 0.893 [0.767–1.000] | 0.455 [0.296–0.615] | 0.634 [0.442–0.768] | 0.281 [-0.148–0.525] | 8.65 [7.17–10.34] |
| NCT | FS4 | Ridge | 0.704 [0.571–0.810] | 0.674 [0.577–0.771] | 0.380 [0.176–0.579] | 0.893 [0.767–1.000] | 0.455 [0.303–0.615] | 0.659 [0.492–0.786] | 0.316 [-0.080–0.551] | 8.54 [7.03–10.01] |
| NCT | FS1 | SVR | 0.696 [0.554–0.800] | 0.671 [0.561–0.768] | 0.363 [0.132–0.556] | 0.857 [0.710–0.966] | 0.485 [0.333–0.657] | 0.609 [0.409–0.750] | 0.234 [-0.202–0.490] | 8.97 [7.43–10.67] |
| NCT | FS3 | Ridge | 0.696 [0.567–0.805] | 0.671 [0.568–0.776] | 0.363 [0.152–0.564] | 0.857 [0.719–0.967] | 0.485 [0.333–0.643] | 0.649 [0.481–0.779] | 0.282 [-0.135–0.532] | 8.89 [7.38–10.38] |

| Outcome | FS | Model | F1 [95% CI] | BalAcc [95% CI] | MCC [95% CI] | Sens [95% CI] | Spec [95% CI] | Pearson r [95% CI] | R² [95% CI] | MAE [95% CI] |
|---|---|---|---|---|---|---|---|---|---|---|
| NCT | FS3 | SVR | 0.687 [0.542–0.800] | 0.668 [0.553–0.772] | 0.349 [0.110–0.558] | 0.821 [0.667–0.952] | 0.515 [0.343–0.684] | 0.609 [0.426–0.757] | 0.192 [-0.268–0.483] | 9.43 [7.87–10.99] |
| NCT | FS2 | Ridge | 0.686 [0.552–0.800] | 0.656 [0.551–0.767] | 0.335 [0.106–0.547] | 0.857 [0.719–0.967] | 0.455 [0.296–0.615] | 0.651 [0.487–0.780] | 0.269 [-0.167–0.535] | 8.91 [7.41–10.47] |
| NCT | FS2 | SVR | 0.676 [0.531–0.785] | 0.653 [0.527–0.758] | 0.321 [0.056–0.532] | 0.821 [0.667–0.952] | 0.485 [0.310–0.656] | 0.617 [0.437–0.761] | 0.171 [-0.312–0.464] | 9.52 [7.94–11.15] |
| NCT | FS4 | XGBoost | 0.656 [0.525–0.776] | 0.648 [0.550–0.765] | 0.299 [0.100–0.526] | 0.750 [0.613–0.917] | 0.545 [0.382–0.697] | 0.484 [0.261–0.675] | 0.146 [-0.225–0.436] | 9.35 [7.59–11.10] |
| NCT | FS1 | RF | 0.627 [0.485–0.750] | 0.602 [0.479–0.724] | 0.212 [-0.046–0.452] | 0.750 [0.586–0.913] | 0.455 [0.286–0.615] | 0.518 [0.290–0.705] | 0.177 [-0.244–0.456] | 9.28 [7.61–11.01] |
| NCT | FS3 | XGBoost | 0.615 [0.483–0.738] | 0.600 [0.482–0.726] | 0.203 [-0.040–0.452] | 0.714 [0.545–0.885] | 0.485 [0.310–0.647] | 0.444 [0.198–0.653] | 0.071 [-0.382–0.378] | 9.84 [8.06–11.63] |

| Outcome | FS | Model | F1 [95% CI] | BalAcc [95% CI] | MCC [95% CI] | Sens [95% CI] | Spec [95% CI] | Pearson r [95% CI] | $R^2$ [95% CI] | MAE [95% CI] |
|---|---|---|---|---|---|---|---|---|---|---|
| NCT | FS4 | RF | 0.615 [0.462–0.746] | 0.600 [0.480–0.724] | 0.203 [-0.043–0.447] | 0.714 [0.543–0.864] | 0.485 [0.310–0.647] | 0.548 [0.324–0.726] | 0.224 [-0.168–0.508] | 8.91 [7.22–10.69] |
| NCT | FS3 | RF | 0.606 [0.475–0.732] | 0.584 [0.471–0.708] | 0.174 [-0.057–0.427] | 0.714 [0.545–0.885] | 0.455 [0.278–0.625] | 0.530 [0.321–0.705] | 0.161 [-0.270–0.462] | 9.41 [7.71–11.05] |
| NCT | FS2 | XGBoost | 0.597 [0.464–0.727] | 0.569 [0.455–0.701] | 0.144 [-0.097–0.404] | 0.714 [0.545–0.885] | 0.424 [0.257–0.590] | 0.462 [0.214–0.669] | 0.070 [-0.408–0.383] | 9.79 [8.01–11.66] |
| NCT | FS2 | RF | 0.594 [0.452–0.727] | 0.582 [0.465–0.706] | 0.166 [-0.073–0.420] | 0.679 [0.500–0.839] | 0.485 [0.321–0.641] | 0.540 [0.333–0.710] | 0.159 [-0.271–0.457] | 9.40 [7.70–11.12] |
| NCT | FS1 | XGBoost | 0.571 [0.414–0.706] | 0.564 [0.435–0.688] | 0.129 [-0.128–0.380] | 0.643 [0.458–0.840] | 0.485 [0.310–0.656] | 0.363 [0.084–0.605] | -0.014 [-0.498–0.316] | 10.03 [8.06–12.22] |

CI = 95% bootstrap confidence interval. Sens = sensitivity; Spec = specificity; PPV = positive predictive value; NPV = negative predictive value.

*Note:* All values are point estimate [lower–upper bound of 95% bootstrap confidence interval] computed from 2,000 resamples of the out-of-fold predictions. Top-performing models (highest F1 per outcome) shown in bold. Models sorted within outcome by F1 (descending). BalAcc = balanced accuracy = (sensitivity + specificity) / 2; MCC = Matthews Correlation Coefficient; Pearson r = correlation between predicted and observed continuous scores; $R^2$ = coefficient of determination; MAE = mean absolute error; CI = 95% bootstrap confidence interval.

*Supplementary Table S3. Complete Feature List by Feature Set.*

| FS | Feature | Type | Description |
|---|---|---|---|
| FS1 | wab_aq_acute | Clinical | Acute WAB-AQ (0–100) |
| FS1 | age_at_stroke | Clinical | Age at stroke onset (years) |
| FS1 | sex | Clinical | Biological sex (M/F) |
| FS1 | education_yrs | Clinical | Years of education |
| FS1 | prior_stroke | Clinical | Prior stroke history (Y/N) |
| FS2 | lesion_volume_mL | Volumetric | Lesion volume in mL |
| FS3 | BPM_IFG_opercularis_L | Cortical | L IFG pars opercularis (BA44) proportional damage |
| FS3 | BPM_IFG_triangularis_L | Cortical | L IFG pars triangularis (BA45) proportional damage |
| FS3 | BPM_STG_L | Cortical | L superior temporal gyrus proportional damage |
| FS3 | BPM_SMG_L | Cortical | L supramarginal gyrus proportional damage |
| FS3 | BPM_AG_L | Cortical | L angular gyrus proportional damage |
| FS3 | BPM_MTG_L | Cortical | L middle temporal gyrus proportional damage |
| FS3 | BPM_ITG_L | Cortical | L inferior temporal gyrus proportional damage |
| FS3 | BPM_Ins_L | Cortical | L insula proportional damage |
| FS3 | BPM_MFG_L | Cortical | L middle frontal gyrus proportional damage |
| FS3 | BPM_SFG_L | Cortical | L superior frontal gyrus proportional damage |
| FS3 | SLF_L_prob | WM | L SLF disconnection probability (dorsal stream) |
| FS3 | SLFt_L_prob | WM | L SLF temporal/arcuate disconnection probability |

| FS | Feature | Type | Description |
|---|---|---|---|
| FS3 | IFOF_L_prob | WM | L IFOF disconnection probability (ventral stream) |
| FS3 | ILF_L_prob | WM | L ILF disconnection probability (ventral stream) |
| FS3 | UF_L_prob | WM | L uncinate fasciculus disconnection probability |
| FS4 | pair_POP-STG | Network | L pars opercularis – L STG |
| FS4 | pair_POP-SMG | Network | L pars opercularis – L SMG |
| FS4 | pair_PTR-STG | Network | L pars triangularis – L STG |
| FS4 | pair_POP-MTG | Network | L pars opercularis – L MTG |
| FS4 | pair_PTR-MTG | Network | L pars triangularis – L MTG |
| FS4 | pair_SMG-STG | Network | L SMG – L STG |
| FS4 | pair_IPG-STG | Network | L IPG – L STG |
| FS4 | pair_ITG-STG | Network | L ITG – L STG |
| FS4 | pair_AG-STG | Network | L AG – L STG |
| FS4 | pair_POP-PTR | Network | L pars opercularis – L pars triangularis |
| FS4 | pair_IN-POP | Network | L insula – L pars opercularis |
| FS4 | pair_IN-STG | Network | L insula – L STG |
| FS4 | pair_SFG-POP | Network | L SFG – L pars opercularis |
| FS4 | pair_L.SMG-R.SMG | Network | L SMG – R SMG (interhemispheric) |
| FS4 | pair_L.STG-R.STG | Network | L STG – R STG (interhemispheric) |
| FS4 | pair_L.POP-R.POP | Network | L pars oper. – R pars oper. (interhemispheric) |
| FS4 | pair_L.PTR-R.PTR | Network | L pars tri. – R pars tri. (interhemispheric) |
| FS4 | pair_L.SFG-R.SFG | Network | L SFG – R SFG (interhemispheric) |
| FS4 | pair_ITG-MTG | Network | L ITG – L MTG |
| FS4 | pair_AG-MTG | Network | L AG – L MTG |
| FS4 | pair_FG-ITG | Network | L fusiform – L ITG |
| FS4 | pair_L.thalamus-POP | Network | L thalamus – L pars opercularis |

*Note*: FS1 = 5; FS2 = 6; FS3 = 21; FS4 = 43 features (43 listed). BPM/Tractotron/NeMo features residualized against lesion volume inside each CV fold.

**Supplemental Figures**

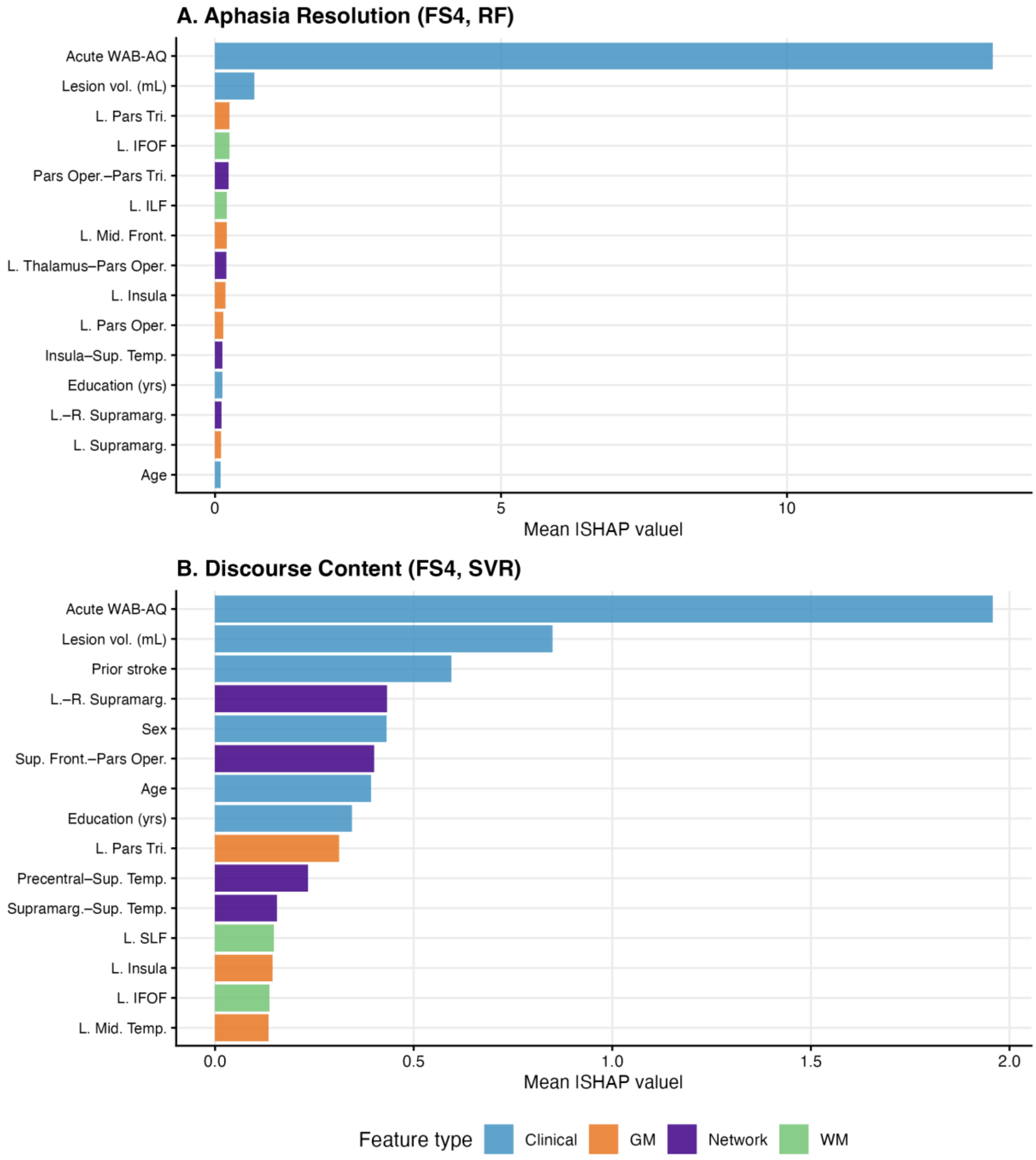


**Supplementary Figure S1. SHAP Bar Summary.** Top 15 features ranked by mean |SHAP| value averaged across all 100 outer test folds for each top-performing model. (A) Aphasia resolution (FS4 RF). (B) Discourse content normalization (FS4 SVR). Bars are coloured by feature modality: clinical/demographic (blue), cortical/BPM (green), white matter/Tractotron (orange), network/NeMo (purple). Note the different x-axis scales between panels.
